# The impact of live dietary microbes on health: a scoping review

**DOI:** 10.1101/2023.08.16.23294168

**Authors:** Ajay Iyer, Arghya Mukherjee, Beatriz Gómez-Sala, Eibhlís M. O’Connor, John G. Kenny, Paul D. Cotter

**Author notes:** Shared first author (Ajay Iyer, Arghya Mukherjee).

## Abstract

**Background:** A systematic approach to collect, peruse and summarise the available information relating to the potential benefits of consuming dietary microbes was pursued in this scoping review. This review focused on the research endpoints, experimental designs, and microbial exposure in experimental as well as observational research work.

**Approach:** Using a structured-set of keywords, scientific databases were systematically searched to retrieve publications reporting outcomes pertaining to the use of dietary-microbes in healthy, non-patient populations. Searches were further tailored to focus on eight different health categories, namely, “antibiotic associated diarrhoea” (AAD), “gastrointestinal health” (GIH), “immunological health” (ImH), “cardiovascular health and metabolic syndrome” (CvHMS), “cancer prevention” (CanPr), “respiratory health” (ReH), “weight management” (WtMgt), and “urogenital health” (UrGH). Quality of evidence available in each publication was assessed using the Jadad scoring system.

**Result:** The search yielded 228 relevant publications describing 282 experimental cases comprising 62 research endpoints overall. A microbial dose of *≥*2 × 10^9^ CFU·day^−1^ was associated with non-negative reported outcomes. Older population groups with a median age of 39 years were associated with positive outcomes.

**Conclusion:** More high-quality research is required investigating the role of dietary microbes in maintaining general health, particularly in the health categories of urogenital health, weight management and cancer prevention.

## 1 Introduction

### Rationale

There are many ways in which humans consume live microbes as part of their diet, in particular, through fermented foods (defined as foods made through desired microbial growth and enzymatic conversions of food components) [1] and through probiotics (defined as live microorganisms that, when administered in adequate amounts, confer a health benefit on the host) [2]. It has been postulated that there may be benefits associated with the consumption of live dietary microbes [3]. However, randomised, double-blinded, placebo controlled trials testing the effects of live dietary microbes are challenging, with the current portfolio of tested microbes and foods being relatively limited. Furthermore, even among the tested food products containing live microbes, the study outcomes, research end-points, methodologies, and quality of evidence is highly diverse. Although critical systematic reviews have previously addressed some specific health outcomes for certain dietary microbes [4, 5, 6, 7], an overarching review of the evidence for health outcomes related to the more general consumption of dietary microbes is warranted, while also having the potential to highlight research gaps for further investigation.

### Objectives

Our objective was to systematically search and collect literature from scientific databases to summarise and generate evidence maps from the extensive body of knowledge relating to the health benefits of consuming dietary microbes. Such a task was contextualised through the research question: “Does consumption of live dietary microbes improve health in typical, non-patient, healthy populations?” [8] Details relating to specific aspects of the question, such as “non-patient” or “consumption” are elaborated below, but generally, the scoping review focuses on the various research endpoints, methods and practices described in published human trials from the perspective of the prophylaxis of lifestyle associated morbidities. Hence, our work attempts to propose a possible adequate intake (AI) value for dietary microbes, similar to a recommended daily allowance (RDA) for live dietary microbes as suggested previously[9, 10]. Notably, such a recommendation does not attempt to address a dietary deficiency, but rather provide an indicator of healthy eating.

## 2 Materials and methods

### Protocol

This work sought to follow the guidelines on structuring the process of scoping review as per the published checklist [11, 12]. The scope and protocol for this scoping review has been published previously [8].

### Search

The National Library of Medicine (MEDLINE), Cochrane Central, and Scopus bibliographic databases were used for systematic literature searching of relevant articles published between 2000 and 2023. Searches were performed to identify literature pertaining to one of eight categories, namely, “antibiotic associated diarrhoea” (AAD), “gastrointestinal health” (GIH), “immunological health” (ImH), “cardiovascular health and metabolic syndrome” (CvHMS), “cancer prevention” (CanPr), “respiratory health” (ReH), “weight management” (WtMgt), and “urogenital health” (UrGH). Further details on the search strategy, terms used and the approach are provided in **Supplementary Text 1.1** and **1.2**.

For the majority of relevant publications, the study endpoints spanned multiple health categories. Thus, the primary category and secondary category corresponded to the main hypothesis explored in the publication. This was generally indicated by the metrics/biomarker used to calculate the experimental sample size. Next, the study outcome itself was categorised as positive, neutral or negative. An outcome was deemed positive when a statistically significant difference in the intervention group relative to its comparator was expected to be beneficial. An outcome was deemed neutral when no significant outcome was noted between the intervention and the comparator group. Lastly, an outcome was deemed negative when the statistically significant change was expected to be detrimental to health.

As an example, consider one of the included publications, Higashikawa *et al*. [13]. The primary category was WtMgt as the sample size calculations were selected based on identifying changes in BMI, body fat mass, and waist circumference (the measurable endpoints). The corresponding outcome was deemed positive as a significant decrease in body fat mass was measured and thus, the “primary outcome” of the study was recorded as positive. The secondary category for this study was CvHMS reflecting the fact that changes in the biomarkers HbA1c and serum lipids were part of an exploratory screening. Since no significant changes were observed for these markers, the “secondary outcome” of this study was neutral.

### Selection criteria

The exclusion criteria have been made available in the published protocol [8]. The details are summarised in Table 1.

**Table 1:** Summary of the criteria for excluding publications.

| Parameter | Exclusions |
| --- | --- |
| Cohorts | Atypical cohorts where the human body is (or expected to be) under duress or exhibiting non-representative physiology, such as patients in critical care (chemotherapy, recurring infections, radiotherapy, emergency ward, etc.), active military personnel posted in war zones, astronauts in training, researchers in expeditions, athletes at all competitive levels, and pregnant, lactating and feeding women. |
| Diseased / non-healthy conditions | Disease conditions recorded in literature such as diabetes, hypertension, hypercholesterolaemia, obesity, irritable bowel syndrome (IBS), inflammatory bowel disease (IBD), lactose intolerance, heart disease, spinal chord injuries, malnourishment, asthma, eczema, kidney stones, gout, small intestine bacterial overgrowth (SIBO), fistulas and gastric bypass surgeries. |
| Adjunctive therapy | Except for AAD, studies describing administration of fermented products in conjunction with other therapeutics, such as cholesterol lowering drugs, weight-loss supplements, calorie-restriction special diets, etc. |
| Non-human subjects | Animal studies and in vitro experiments |
| Non oral administration | Delivery of live microbes topically, rectally, nasally, vaginally, etc. |
| Non-viable microbes | Killed-microbes, microbial cell-wall, or specific cell wall proteins as vaccines |
| Non clinical outcomes | Studies reporting solely on the colonisation of microbes in the gut, change in gut microbial compositions, etc. |
| Non experimental literature | Reviews, commentaries, lectures, letters to Editor, presentations, book chapters, posters |
| Confounders | Additives to the dietary microbe containing product that may significantly change or modulate the research endpoint. This included vaccinations, antibiotics for treatment or prophylaxis (except AAD where antibiotics were used for treatment), phenolics or plant bioactives. Non-sterile placebos (presence of *Streptococcus thermophilus* and *Lactobacillus delbrueckii* subsp. *bulgaricus* commonly used in yogurt production) or placebos containing prebiotic (such as fructo-oligosaccharides) |
| Anthropometrics | Age: <2 yr., Body mass index (BMI): $\geq 30 \text{ kg}\cdot\text{m}^{-2}$, Diastolic and systolic blood pressures: $\geq 80 \text{ mm}\cdot\text{Hg}$ and $\geq 130 \text{ mm}\cdot\text{Hg}$, respectively, HbA1c: $\geq 6.1\%$, LDL, total triglyceride and total cholesterol levels: $\geq 135 \text{ mg}\cdot\text{dL}^{-1}$ ($3.50 \text{ mmol}\cdot\text{L}^{-1}$) and $\geq 170 \text{ mg}\cdot\text{dL}^{-1}$ ($4.4 \text{ mmol}\cdot\text{L}^{-1}$), and $\geq 230 \text{ mg}\cdot\text{dL}^{-1}$ ($6.0 \text{ mmol}\cdot\text{L}^{-1}$), respectively., Nugent scores: $\geq 6.$, Fasting glucose levels: $\geq 100 \text{ mg}\cdot\text{dL}^{-1}$ ($5.6 \text{ mmol}\cdot\text{L}^{-1}$), Fasting insulin level: $\leq 2 \mu\text{IU}\cdot\text{mL}$ and $\geq 6 \mu\text{IU}\cdot\text{mL}$. |

### Data items

The following information was extracted from the literature and provided in a master-table as a .csv file (Supplementary Table 1): year of publication, aim of the paper, country of the research institution(s), registration numbers (where available), research category as defined in this study (e.g., AAD, respiratory health), blinding status, randomisation status, placebo components (if used), study design (parallel, cross-over, single-arm, etc.), the food or supplement tested, status of the microbe (freeze dried, spores, etc.), dietary microbes provided in the study, microbial dose per unit test product, number of doses in a day, duration of active microbe consumption (in days), overall duration of study (in days), volunteer numbers in experimental and placebo groups, males recruited in the volunteer group (%), age of volunteers (in years), body mass index (BMI) of volunteers (in kg·m^−2^), target population, exclusion criteria of the study, primary outcome, secondary outcome, adverse events (when reported), study flow (study visits and sampling periods), result notes, funding body and conflict of interest, statistical methods used (sample size calculation, statistical tests, software used, per protocol or intention to treat (ITT), etc), recruitment success (number of people who participated in the study against those who were screened, expressed in %), remainers (volunteers who successfully adhered to study guidelines and completed the study, expressed in %), Jadad score (if the publication was not a population/observational study), disparity between sample size requirement and final study participation, method used for compliance check, compliance value (%)

### Synthesis of results

Publications with multiple studies: Some publications reported multiple studies to demonstrate the overall effect of products containing live dietary microbes. In such instances, the studies were examined individually. For example, Östman *et al*. reported [14] two studies to demonstrate the general benefits of fermented foods, where dairy products (filmjölk or långfil) were investigated in the first, and fermented vegetables (pickled cucumber) in the second. Each study had its own controls and was individually assessed and further split into “cases” as elaborated below.

### Study with multiple cases

In some publications, multiple products were compared against a control (e.g.: Actimel versus “dahi” versus sterile yogurt [15]) or a dose response was investigated (10^8^ versus 10^9^ versus 10^10^ versus 10^11^ CFU·mL^−1^ versus placebo [16]). While the basic parameters of the setup were the same, the treatments (either dose or duration or the composition of the dietary microbe-containing product) were different. Thus, the studies were further divided into “cases”, which accounted for each variation in the treatment. Statistically, each case was treated as a unique data point obtained through independent experimentation. This assumption was based on the randomisation and division of volunteers into each group or treatment and the general trend that grouping was proportionally representative.

### Microbial nomenclature and dosage uncertainties

Since the time period included in this study ranged from the year 2000 to 2023, many publications have used outdated taxonomic names for microbes found in probiotic and fermented foods. Many recent publications also continue to use outdated nomenclature, particularly when the test product is a branded probiotic. The list of all reported bacterial strains along with their corresponding updated names (as on 8 February 2023) is provided in **Supplementary Table 2**. This scoping review uses the updated names of the microbes.

In many publications, the dosages of the constituent bacterial species was often expressed as a total CFU count. In such cases, in the absence of further information, it was assumed that the CFU count of each bacteria was equal. For example, in a publication by Rajkumar *et al*. [17] the concentrations of Streptococcus thermophilus and Lactobacillus bulgaricus was collectively expressed as 10^6^ CFU·mL^−1^. Thus, when considering the expected concentration of each strain, the concentration of each of the two bacterial species was assumed to be 5 × 10^5^ CFU·mL^−1^.

### 2.1 Data visualisation

Graphing and data-wrangling were performed with R (version: 4.1.2) [18] using the tidyverse package [19].

## 3 Results

### 3.1 Literature recovery

The database search yielded 228 publications relevant to the search specifications. Among the 228 publications, all except one [20] had digital object identifiers or PubMed Central IDs which allowed for reliable digital document tractability. However, only 100 publications had any form of registration either in their local government or public registries such as clinicaltrials.gov [21]. Thus, subsequent follow-up or study updates required more effort to trace. The PRISMA-ScR flow-chart of the present scoping review is provided in Figure 1 below:

**Figure 1:**
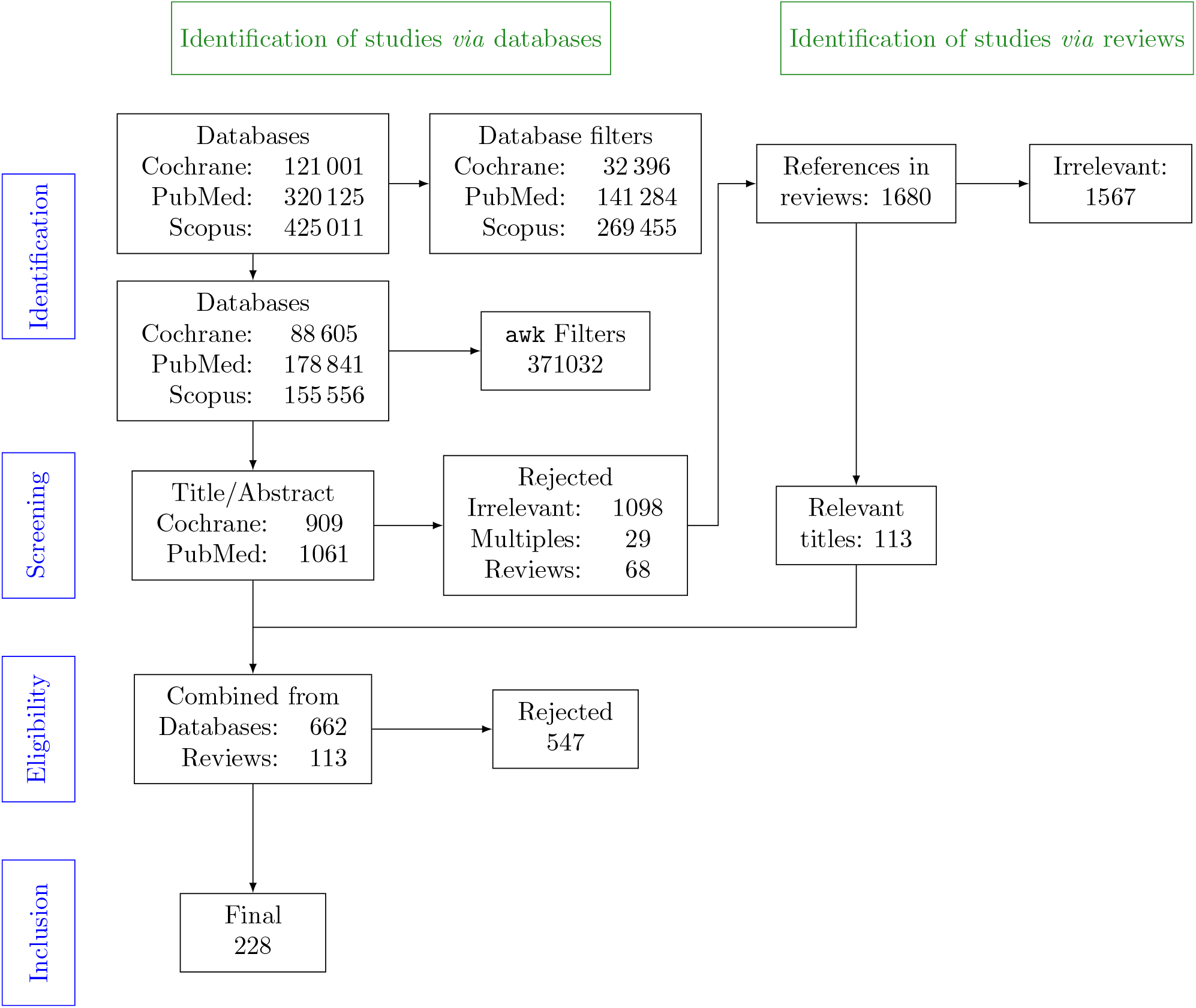
The PRISMA-ScR flowchart indicating the databases used, the methods used to filter results, and the final inclusion of publications.

The publication counts per year are provided in **Supplementary Figure 1**. A loess-smoothened curve was fit to gauge the overall trend in publication. Relevant publications peaked between 2016 to 2017.

Across the 228 publications, these studies could be further divided into 282 cases which differed by the setting (hospital setting versus community setting [15]), or co-reporting two independently performed studies [22]. The information summarised in the following sections may be grouped either at the publication level (n=228), or case level (n=282) depending on the context and relevance to the discussion.

### 3.2 Publication summary

A summary of the various findings at the publication level (n=228) in the literature is provided in Table 2 below. A consolidated master table with the greater details of each study is provided in **Supplementary Table 1**.

**Table 2:**
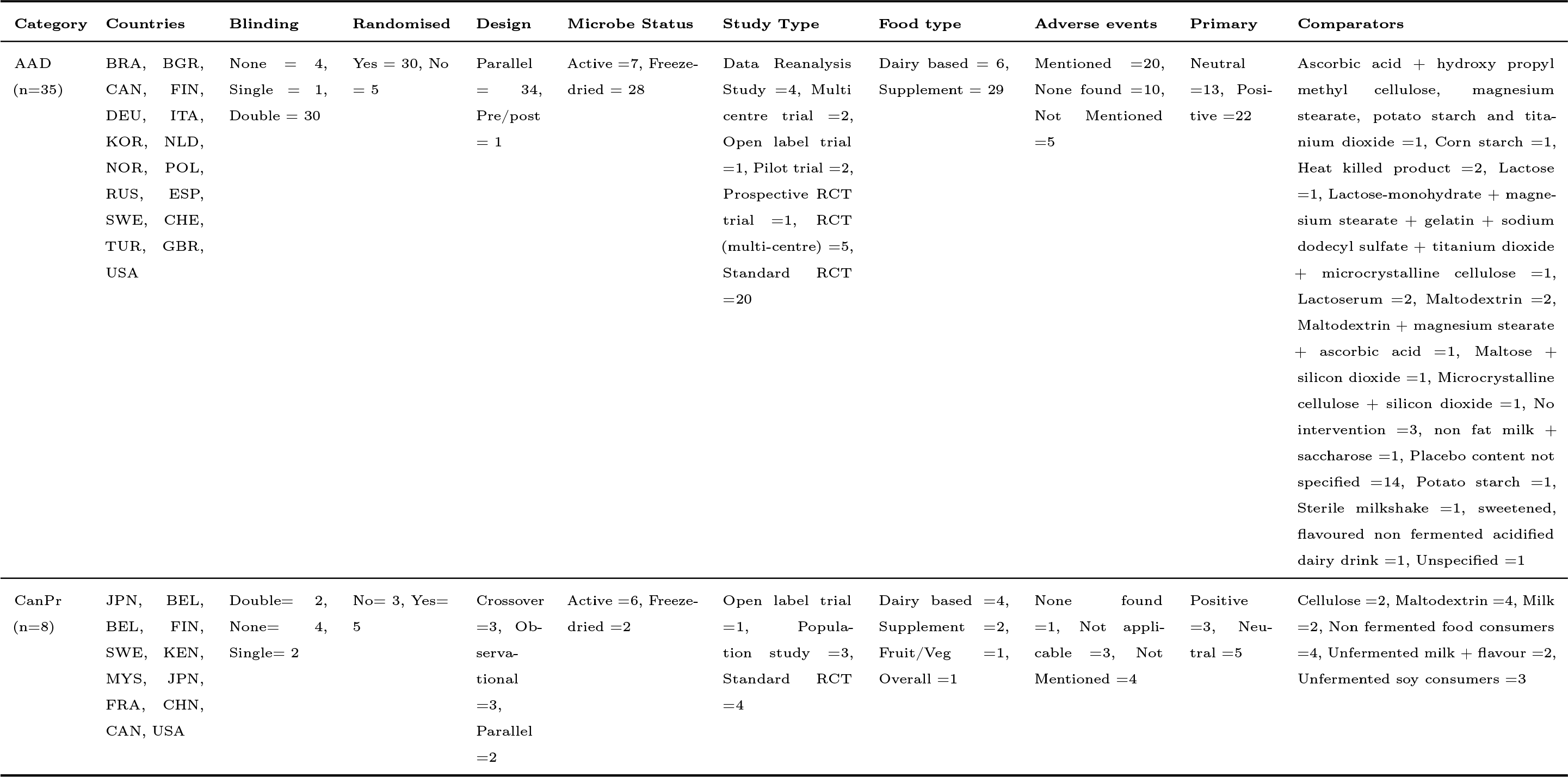
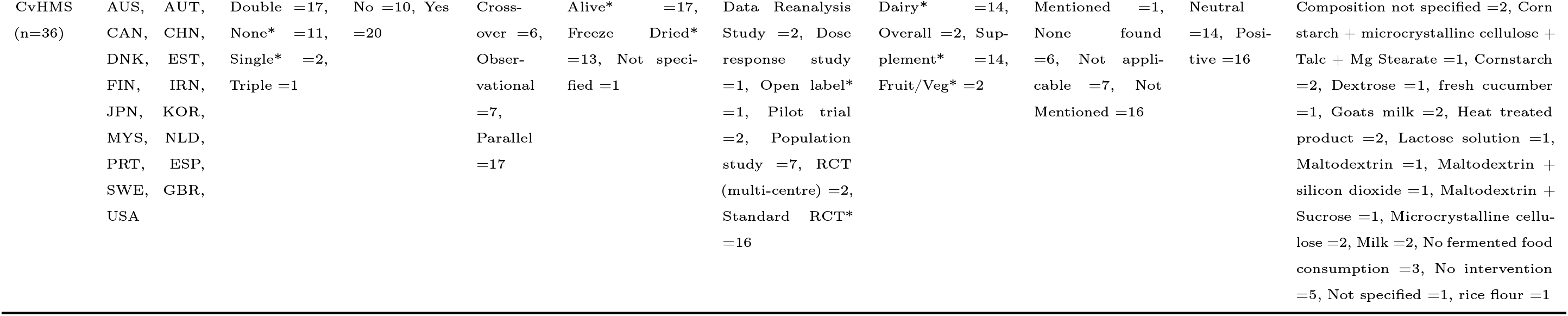
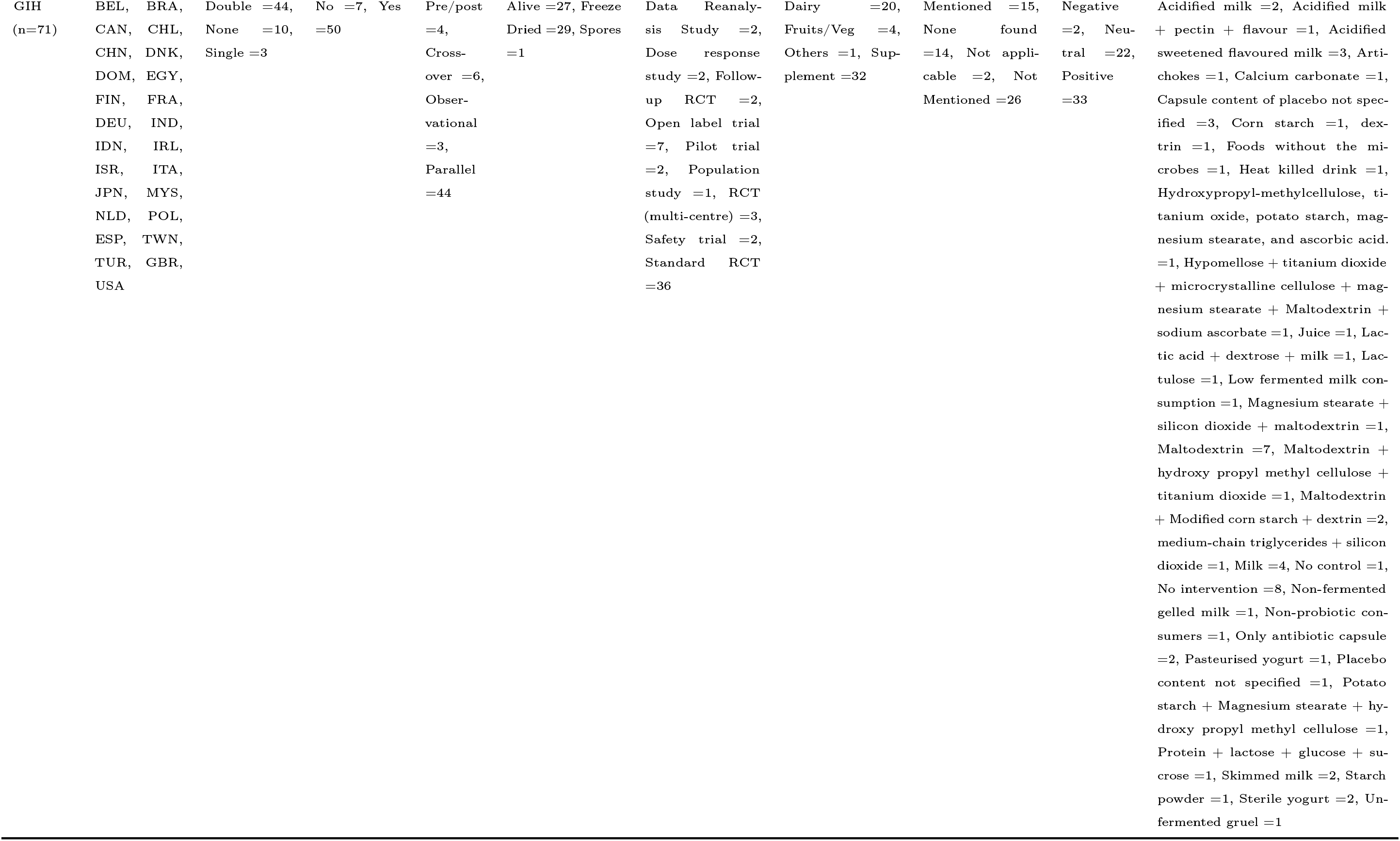
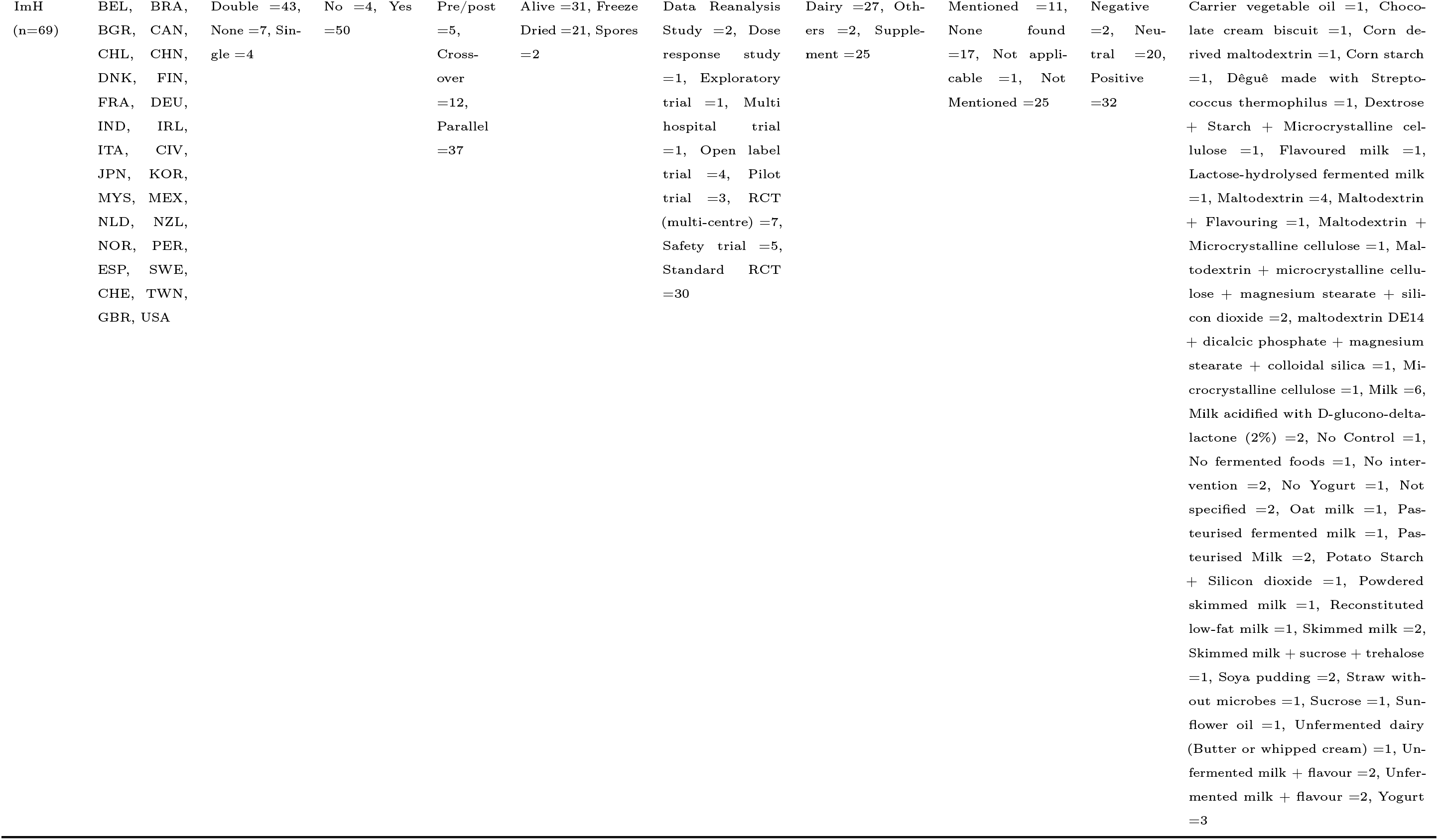
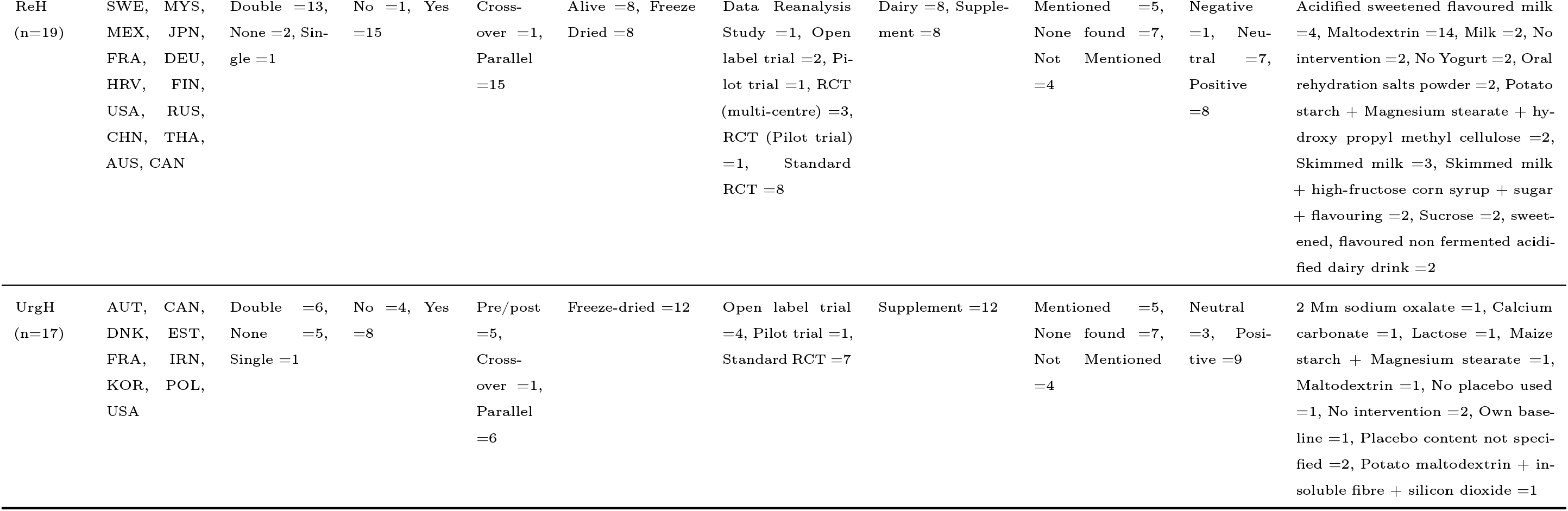
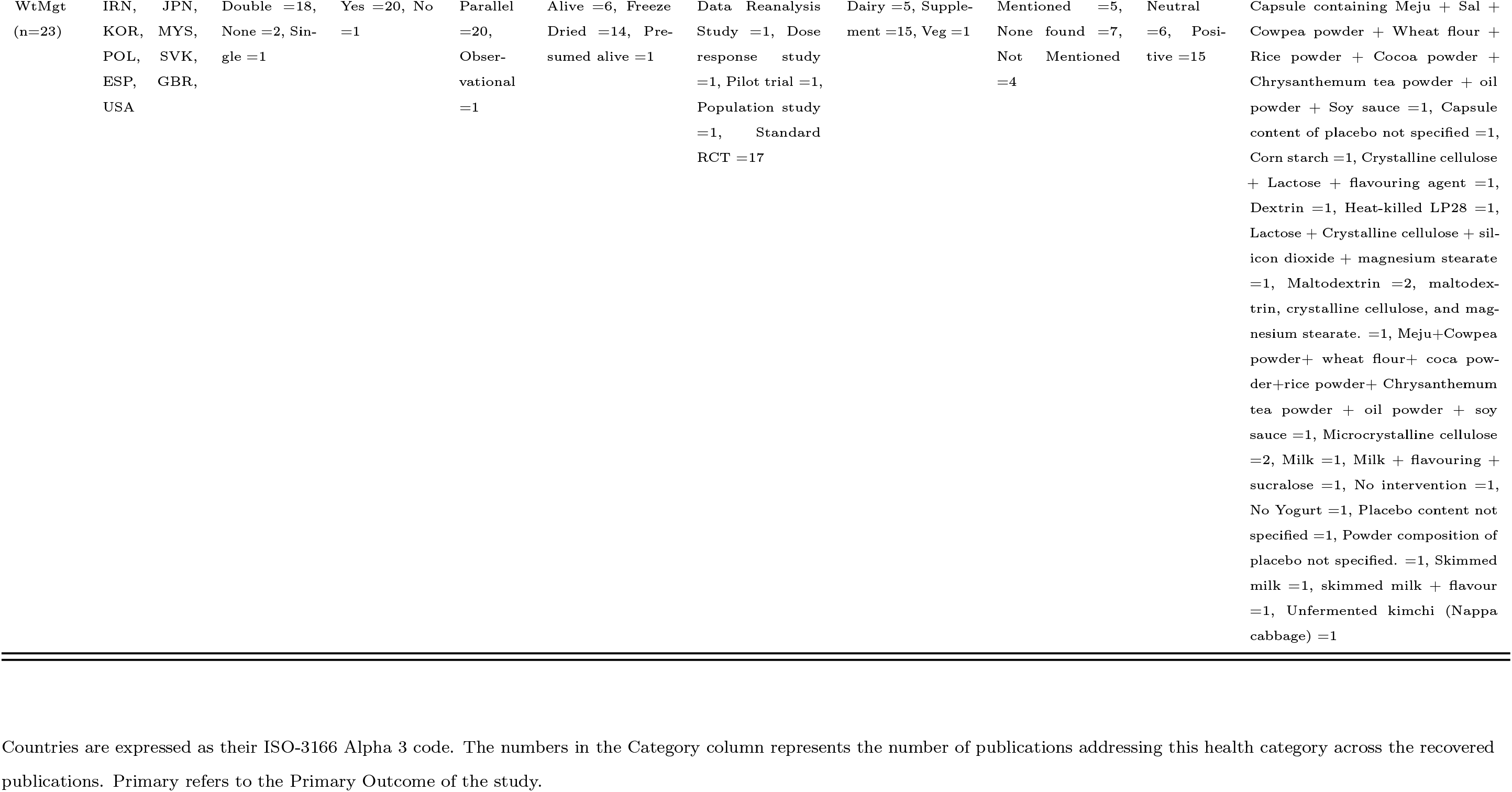
Summary of the study features for each health category from the 228 recovered publications.

### 3.3 Health categories across studies

Across 228 publications, 35 related to antibiotic associated diarrhoea / clostridium associated diarrhoea (AAD), 8 related to cancer prevention (CanPr), 36 related to cardiovascular and metabolic health syndrome (CvHMS), 71 related to gastrointestinal health (GIH), 69 related to Immunological health (ImH), 19 related to respiratory health (ReH), 17 in urogenital health (UrGH), and 23 related to weight management (WtMgt), were recovered.

Finer resolution of the data could be observed when considering cases (n=282), as it was possible to visualise instances where the same health category was represented in the primary as well as the secondary research endpoint. This is depicted in Figure 2.

**Figure 2:**
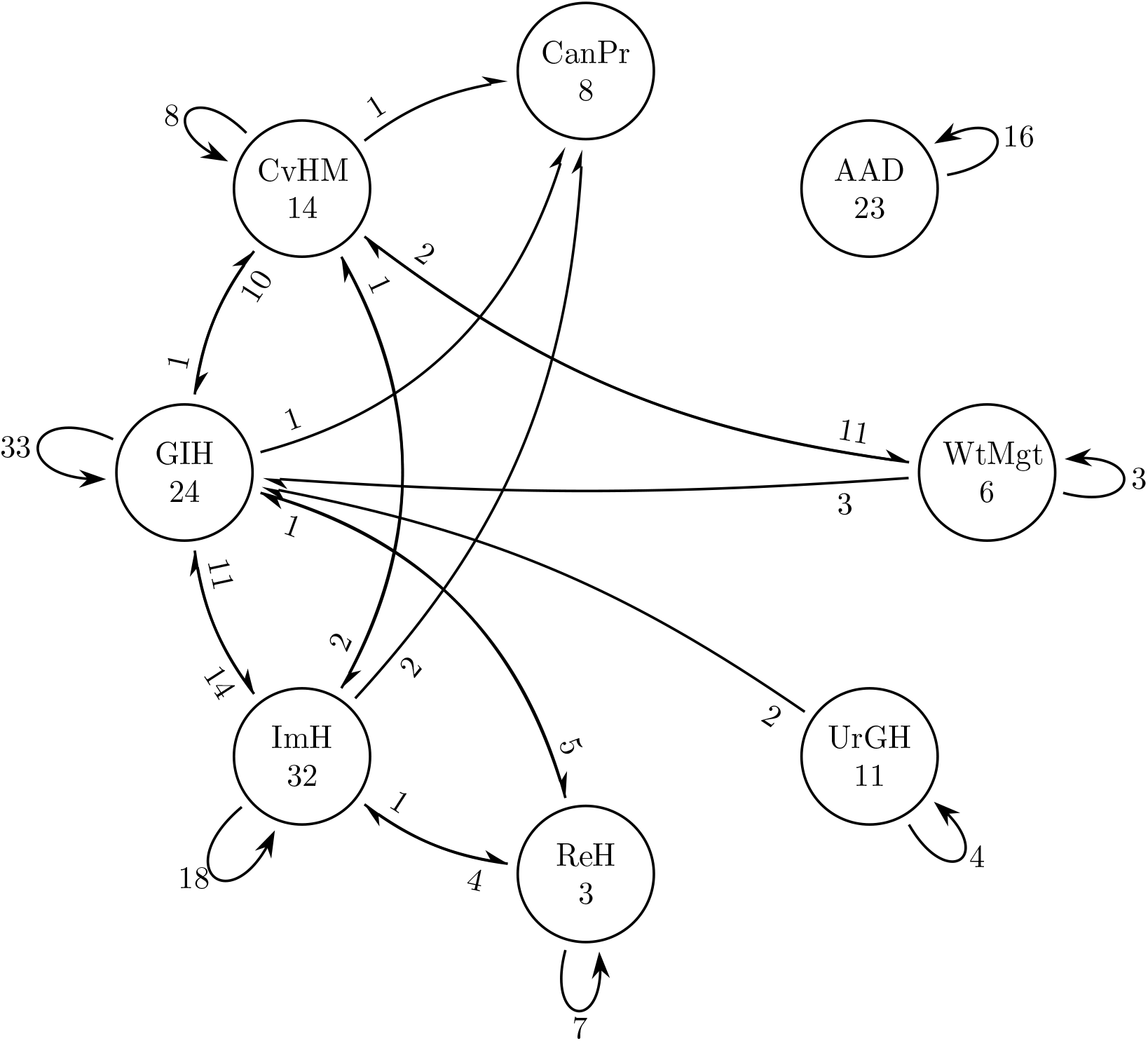
Connection graph of the health categories represented in each case. The number within the node represents the number of times a category was represented as the primary and only category. The loop represents the number of times a category was the primary as well as secondary reported category. In the arrow connecting the nodes, the number proximate to the node represents the number of times the corresponding health category was represented as primary research end-point, while the health category to which the arrow leads represents the secondary health category for that research endpoint.

Nineteen cases where the secondary outcome was a change in the gut microbial composition were excluded from the counts for gastrointestinal health as modifications of gut microbial composition were not regarded as a relevant clinical outcomes.

Figure 2 helps to illustrate potential gaps in live dietary microbe research that link aspects of human physiology. Categories that were unlinked in the literature were WtMgt and CanPr, UrGH and CanPr, ReH and CanPr, ReH and CvHMS, UrGH and CvHMS, UrGH and WtMgt, ImH, and WtMgt. AAD may be considered as a specialised case of gastrointestinal health (GIH).

### 3.4 Study characteristics

Participation: Across the 228 cases, a cumulative number of 299,635 participants were noted, with a median of 72 participants in each publication. The largest number of participants, n=75,089, took part in an observational study [23], while the lowest participant number was n=8 [24].

When grouped according to study primary categories, cumulative participation in studies relating to AAD was n=19,021, CanPr was n=156,708, CvHMS was n=87,156, GIH was n=13,153, ImH was n=5298, ReH was n=7529, UrGH was n=554 and WtMgt was n=10,216. The median, minimum and maximum participation in each category (respectively) was n=147, n=23, and n=5093 for AAD, n=74.5, n=10, and n=75,089 for CanPr, n=6535, n=14, and n=38,802 for CvHMS, n=61, n=12, and n=6548 for GIH, n=50, n=8, and n=638 for ImH, n=368, n=80, and n=1783 for ReH, n=50, n=11, and n=76 for UrGH, and n=80, n=23, and n=8516 for WtMgt.

#### Experimental Designs

Various experimental study designs were noted across the study cases which could be broadly classified into: pre/post (n=20), parallel (n=212) and crossover (n=31), with the remaining n=19 being observational and follow-up studies. One study [25] that initially employed a crossover design, switched to a parallel design when carry-over effects were noted during compliance checks.

#### Microbial status

The products containing dietary microbes reported in the literature can be broadly categorised into two classes based on their expected metabolic status, either being metabolically active (such as in fermented foods like yogurt, kimchi, etc.), or metabolically inactive (such as in freeze–dried or dessicated supplements) or spores at the time of consumption. Spores were considered as a separate category because they were metabolically inactive through a unique biological process, and are physiologically different from freeze-dried vegetative cells.

Across the 282 cases, there were 126 cases where the live dietary microbes were expected to be metabolically active in the food products. The microbe status was freeze-dried/lyophilised in 152 cases, and lastly in 3 cases, the microbes were delivered as spores. There was one case where the status of the microbe was neither specified nor could be discerned from the experimental details. Among the cases where the microbes were metabolically active, 4 reported negative outcomes, 56 reported neutral outcomes and 66 reported positive outcome. For freeze-dried live dietary microbes, 2 cases reported negative outcomes, 60 reported neutral outcomes, and 90 reported positive outcomes. Lastly, for microbes as spores, 2 reported negative outcomes and one reported positive outcomes.

#### Food status

Across 282 cases, the consumed products were broadly classified as either fermented foods (n=31) or as probiotic products (n=251). In cases where microbes were separately added to a food product, they were placed in the probiotic category as these microbes were not part of the fermentation step required for food preparation. Among the publications investigating fermented foods, 14 had neutral outcomes, while 17 had positive outcome. In publications investigating probiotic products, 6 reported negative outcomes, 104 reported neutral outcomes, and 141 reported positive outcomes. The number of studies focusing on fermented foods was generally low, lending credence to the concerns raised in the consensus statement on fermented foods by Marco *et al* regarding the representation of fermented foods in research [1].

#### Strain or species numbers

The number of unique strains of dietary microbes noted in a food product was used as an indicator of its constituent microbial diversity. There is a difference of opinion on whether or not microbial diversity in food products has significant bearing on the health outcomes. While Ouwehand *et al*. [26] showed a significant positive effect, Ritchie and Romanuk [27] could not find any significant association. The foods from the collected literature were divided into 1, 2, 3 and >3 constituent strain numbers, and corresponding counts in the 282 cases, and the associated doses (CFU·day^−1^) were noted (Figure 3). Note that in cases where fermented products had undefined starters, they were assumed to be >3 as they were expected to have a composite microbiome that not only contributed to the fermentation process and food quality, but was also being actively consumed.

**Figure 3:**
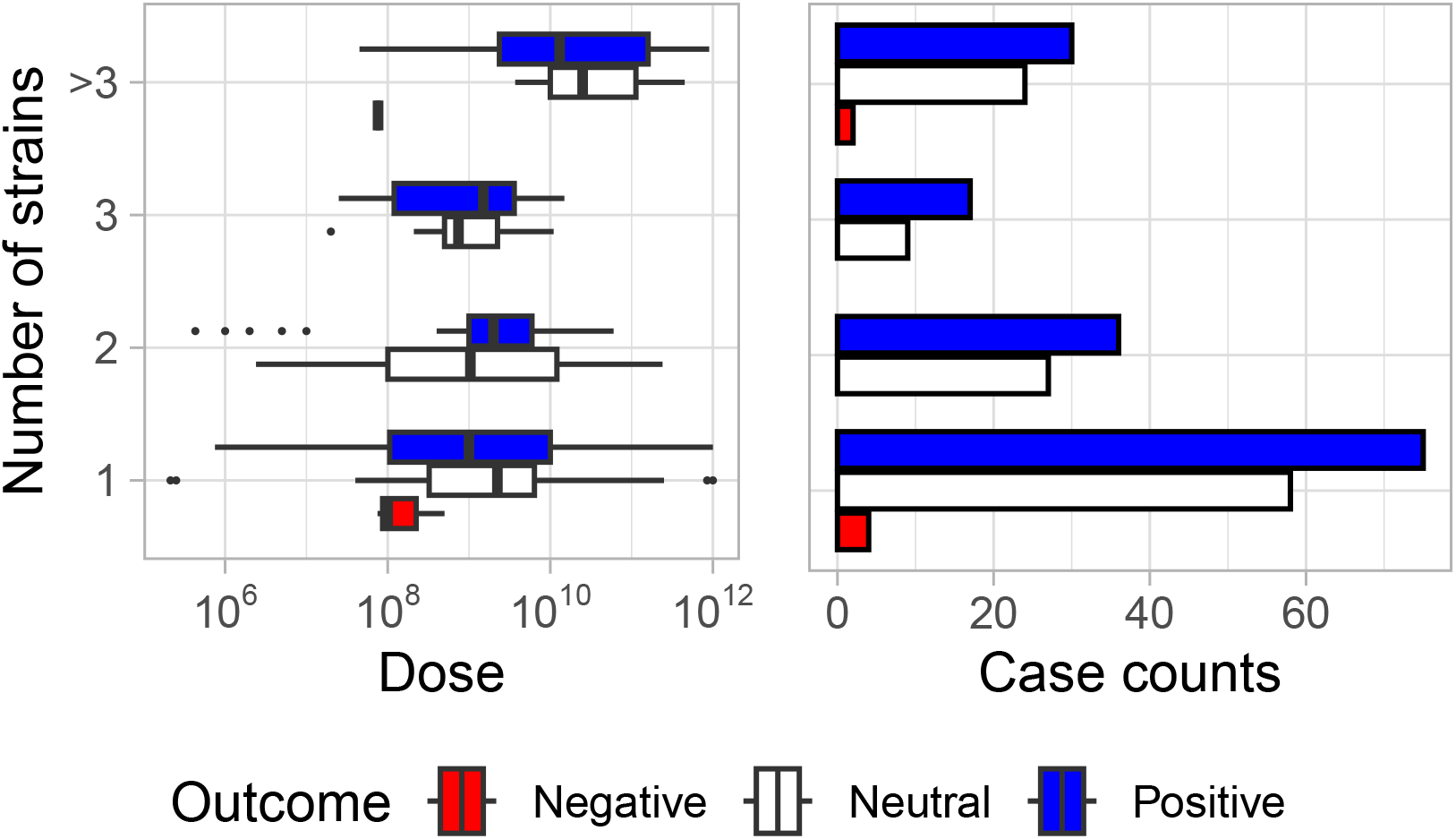
Counts and corresponding doses (CFU·day^−1^) grouped by outcome for foods with 1, 2, 3, or >3 constituent microbial strains. Panel on the left shows the overall microbial dose noted in the food product. Panel on the right shows the case counts (frequency) of food products noted in literature with 1, 2, 3, or >3 constituent microbes.

Notably, some negative outcomes were noted in studies where the food product contained 1 or >3 constituent microbial strains (Figure 3). While the general grouping of experimental microbial doses administered across the literature is described in detail in the following sections, lower doses were associated with negative outcome.

### 3.5 Study population characteristics

The common populations demographics reported across the cases for all health categories were participation of sexes in the study expressed as % males, body mass index (BMI, kg·m^−2^), and the age (y). The overall distribution of the population data grouped by outcome is shown in **Supplementary Figure 2**. The median age of the positive, neutral and negative outcome groups 39.0 y, 32.2 y and 31.4 y respectively. Figure 4. shows trends in sex, age and BMI for all health categories.

**Figure 4:**
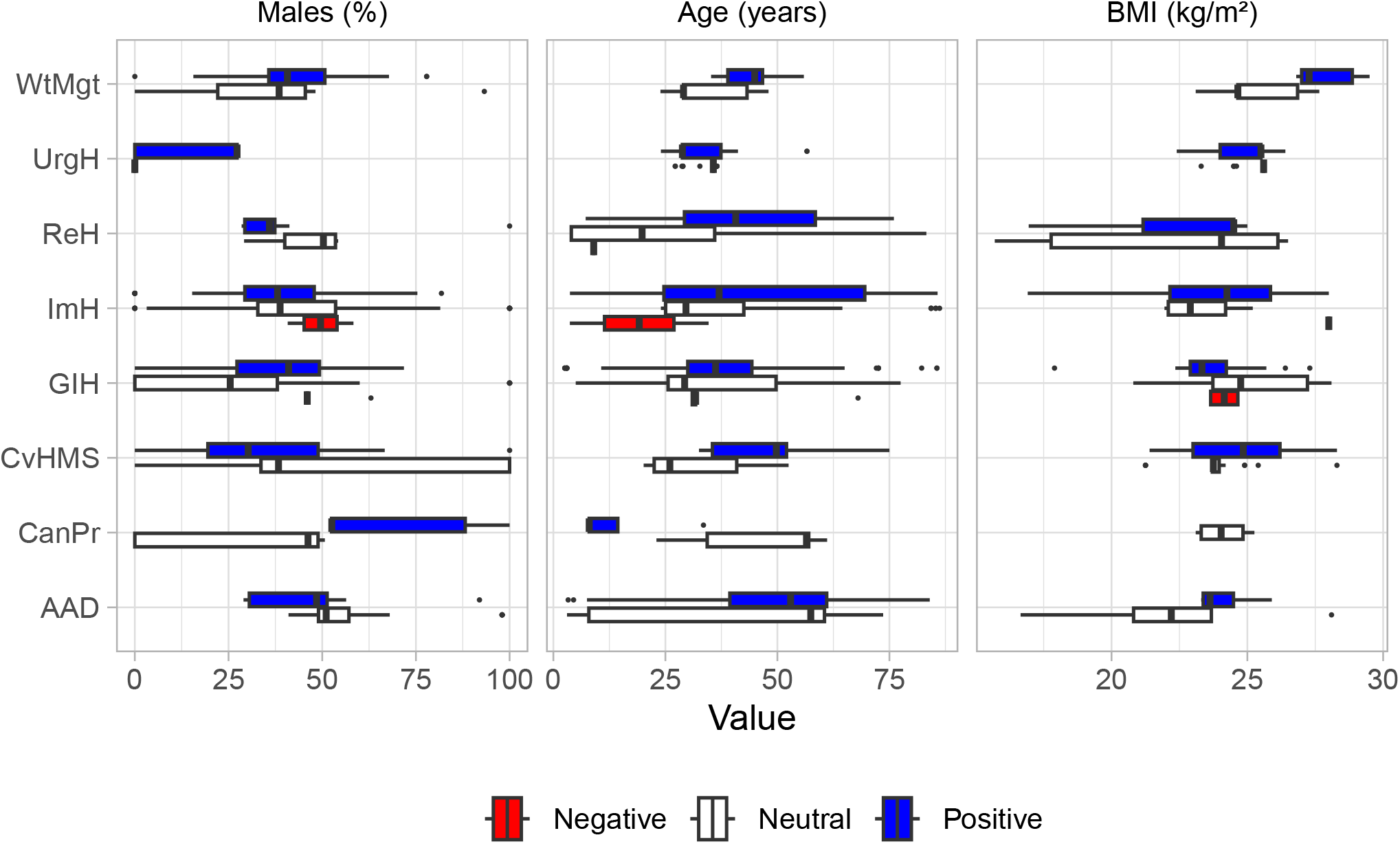
Boxplot depicting the distribution of the population demographics, namely, males (%) representing the percentage of men in the study participation, age (years) representing the mean age of the study participants, and BMI (kg·m^−2^) across the eight health category. Boxplots are further sub-grouped to show distribution of the population parameters for each outcome (positive, neutral or negative).

The secondary outcomes are shown in **Supplementary Figure 3**.

### 3.6 Live dietary microbes intake trends

Overall live microbe intake: The experimental effects of live dietary microbes were assessed on three major parameters, namely, the daily dose (log_10_ transformed, CFU·day^−1^), the duration of experimental intervention (day) and the total exposure to dietary microbes (log_10_ transformed, CFU). Total exposure was calculated as the product of the untransformed daily dose and experimental intervention duration, and subsequently log_10_ transformed.

For positive, neutral and negative primary outcomes, the corresponding median dose of dietary microbes was 2 × 10^9^ CFU·day^−1^,3 × 10^9^ CFU·day^−1^ and 7.6 × 10^7^ CFU·day^−1^ respectively. The median duration of studies for positive, neutral and negative outcomes were 28 day, 28 day and 14 day respectively. For positive, neutral and negative primary outcomes, the corresponding median exposure to live dietary microbes was 2.8 × 10^11^ CFU, 2.8 × 10^11^ CFU and 3.2 × 10^9^ CFU, respectively. The overall comparison of dose, study duration and total exposure stratified by outcome is shown in **Supplementary Figure 4**.

When bifurcating the data based on whether dietary microbes were consumed in fermented foods or probiotic products, the median dose for neutral and positive outcomes was: 8 × 10^8^ CFU·day^−1^ and 5.5 × 10^8^ CFU·day^−1^ respectively for fermented foods, and, 7.6 × 10^7^ CFU·day^−1^, 3 × 10^9^ CFU·day^−1^ and 2 × 10^9^ CFU·day^−1^ for negative, neutral and positive outcomes respectively for probiotic products. The median duration for neutral and positive outcomes were: 10 day and 28 day respectively for fermented foods, and 14 day, 28 day and 28 day, for negative, neutral and positive outcomes respectively for probiotics. Lastly, median exposure for neutral and positive outcomes was: 4.5 × 10^11^ CFU and 1.2 × 10^11^ CFU, respectively for fermented foods, and 3.2 × 10^9^ CFU, 2.7 × 10^11^ CFU and 2.8 × 10^11^ CFU, respectively for probiotics. As noted in section 3.4 above, the number of studies focusing on fermented foods was limited. Thus while being informative, the median values presented separately for fermented foods has limited potential in indicating general trends.

Distribution of data of microbial dose, duration of consumption, and overall exposure to microbes, for each outcome (positive, neutral, or negative) is shown in Figure 5, according the health categories and the food type.

**Figure 5:**
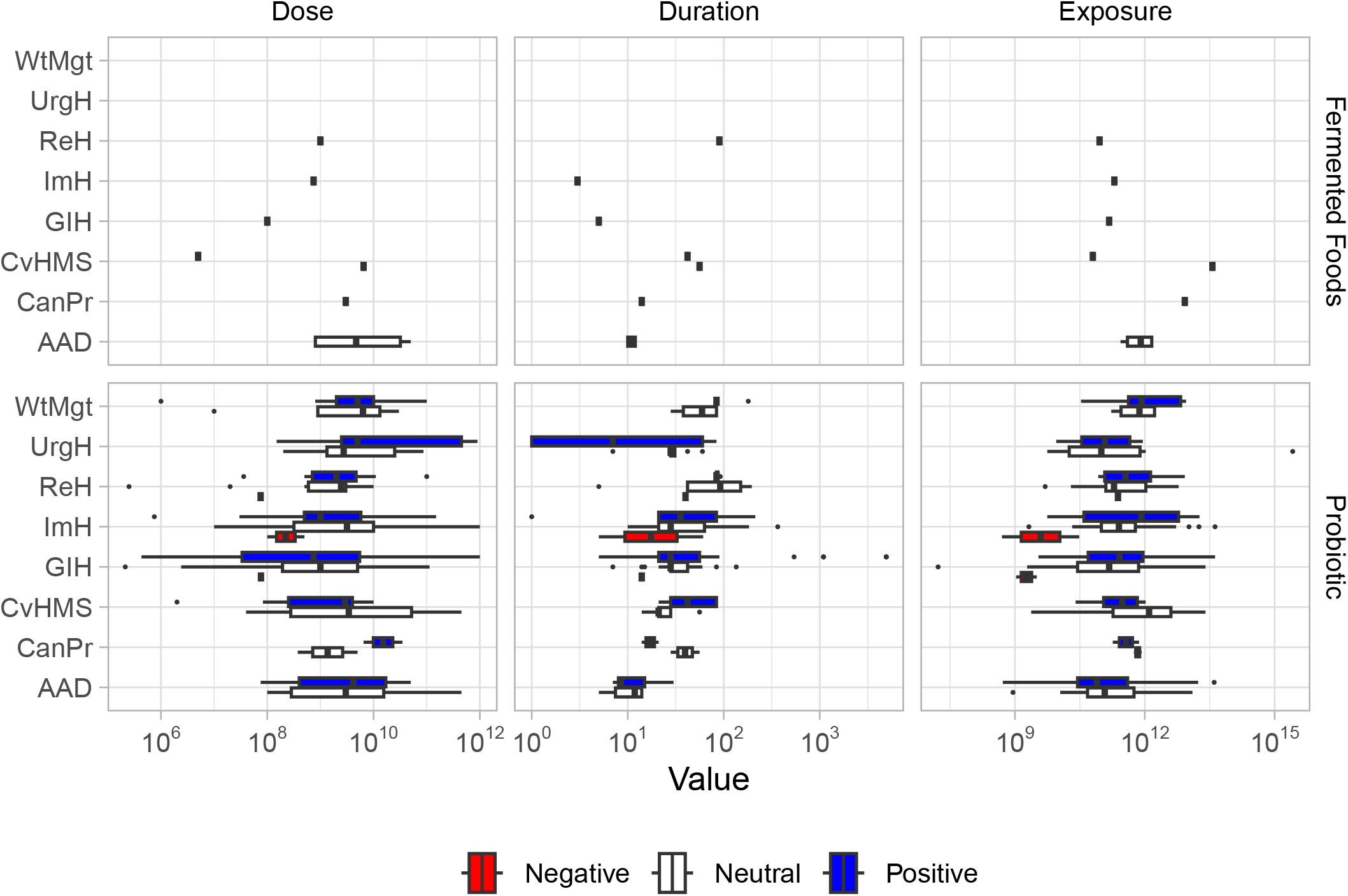
The study dose, duration and overall exposure to dietary microbes grouped according the study outcome for each health category and faceted by the reported types (Fermented foods or Probiotic). Dose represents the microbial dose administered to the study participants. Duration represents the duration of the study where the fermented food containing dietary microbes was actively consumed. Exposure represents the product of Dose and Duration, and represents the total exposure / consumption of the dietary microbes by the study participant.

Corresponding trends for secondary outcomes are shown in **Supplementary Figure 5**.

#### Possible Adequate Intake (AI) value

One of the aims of this work was to assess patterns in the data that may suggest a daily AI dose of live dietary microbes. Figure 5 and **Supplementary Figure 4** appear to suggest that lower doses and low overall exposure to live dietary microbes was associated with a negative outcome. When considering cases with neutral and positive outcomes, the median dose was 2 × 10^9^ CFU·day^−1^. This suggests that this intake value may be recommended as the dose to avoid negative health outcomes.

The median dose values noted for negative, neutral and positive outcomes appear to be supported by the NIH factsheet stating that higher exposure to live dietary microbes does not necessarily translate to positive outcomes [28], although as our findings suggest, may make negative outcomes less likely.

Further high-quality, dose-based studies in tandem with meticulous meta-analyses is required to make robust intake recommendations. Moreover, longitudinal studies are of paramount significance.

#### Microbes stratified by genus

The strain names obtained from the literature were checked and assigned on the basis of the latest taxonomic classification (as on 8 February 2023). Across the recovered literature, a total of 43 unique strains and 23 unique genera were found. In one case relating to kimchi [29], the total *Lactobacillus* count was provided (*Lactobacillus* spp.), which was included in the genus level analyses.

The following 11 genera appeared fewer than six times across the literature, namely, *Alkalihalobacillus* (n=1), *Bacteriodes* (n=4), *Clostridium* (n=1), *Lactoplantibacillus* (n=3), *Latilactobacillus* (n=1), *Lentilactobacillus* (n=2), *Leuconostoc* (n=2), *Levilactobacillus* (n=3), *Pediococcus* (n=2), *Propionibacterium* (n=3), and *Weissella* (n=1). The reported dose (CFU·day^−1^) counts of the remaining 12 genera are depicted in **Supplementary Figure 6**. Among the 12 genera, six reported cases with neutral or positive outcomes. For *Streptococcus*, *Limosilactobacillus*, *Lactobacillus* and *Lacticaseibacillus*, some cases showed a negative primary outcomes. Similarly, *Lactobacillus* sp., were noted to have some negative secondary outcomes.

### 3.7 Statistics applied in the literature

#### ITT versus per protocol

Across the total number of cases 282, the intention-to-treat (ITT) analysis was performed on 49 cases, while 220 cases either directly mention per-protocol or imply so through their data analysis. Cases of observational and population studies (n=13) do not apply here.

#### Statistical tests and practices

In the 282 cases, a variety of a priori tests were employed to evaluate adherence to the assumptions for the parametric tests, either individually or in conjunction with other tests. They were F-test (n=1), *χ*^2^ goodness-of-fit (n=1), Kolmogorov-Smirnov’s test (n=12), Levene’s test (n=6), Shapiro-Wilk’s test (n=19), Z-score transformation (n=3), visual inspection of histograms (n=1), Q-Q residual plots (n=1), logarithmic transformation of data (either log_2_ or log_10_, n=13). In one case [30] statistical methods were unspecified. In six cases, the authors mention the use of either a parametric or non-parametric test based on the measured normality, but do not specify the test used. Lastly, there were 10 observational experiments where no explicit mention of the normality tests were made. In the remaining 223 cases, there was no clear mention of any tests used to check if the data satisfied the assumptions for parametric test.

Across the methods used for statistical comparison/hypothesis testing, 110 cases reported using only parametric tests, while 77 cases reported the use of only non-parametric tests. There were 71 cases where both methods were used as appropriate. In four cases, regression models were used, but the model type was not specified. As noted before, for one case, no information was specified [31], while in 8 cases, the actual statistical test used was not performed and comparisons were descriptive and qualitative with either the display of dimension reduction models or simple expression of means without further analysis. In one case, an HP-nomogram model was constructed that does fit any specific criteria (although may be considered as a regression model as per some definitions of the method). Lastly, the 10 observational studies used Cox-proportional models that are considered semi-parametric (can be loosely considered a special case of the Poisson distribution of factor variance [32].

Lastly, with respect to the use of post hoc corrections and tests to check for model validity and account for biases; across the cases, Benjamini-Hochberg correction (n=2), Bonferroni correction (n=15), Greenhouse-Geisser adjustment (n=2), Dunn’s test (n=2), Dunnett’s test (n=8), Fisher’s Least Significant Difference (n=4), Friedman’s test (n=1), Huynh-Feldt correction (n=1), inverse probability weighting (n=1), Kenward-Roger method (n=4), Mantel-Hanszel test (n=1), McNemar test (n=2), Šidák test (n=1), Steel-Dwass test (n=1), Tukey-Kramer test (n=19), and Yates’ correction (n=2) were used either singly or in combination. Residual analysis was used once in human trials and for testing the validity in all the 10 observational studies and, thus, was reported in 11 cases overall. In one case [33], *post hoc* adjustment was mentioned, but the method was not specified. Thus, ignoring the case where no detail relating to statistical tests was provided, 208 cases did not mention any use of *post hoc* tests.

#### Proprietary versus libre/open-source software

The software used for statistical analyses can greatly influence the scope for reproducibility, largely via the ability to share code. Libre software such as R allow for code-sharing and help with transparency and auditing. Across the 282 publications, only 11 (3.9%) used libre software, while 165 (58.5%) exclusively used proprietary software. Only 9 (3.2%) publications used a mix of the two software types, while the remaining 97 (34.4%) did not detail the software used for statistical analyses. Details are depicted in **Supplementary Figure 7**.

### 3.8 Quality of evidence

#### Quality assessment

The Jadad scoring system, ranging from 0 (very poor) to 5 (rigorous), was utilized to assess the quality of the clinical trials in terms of their adherence to randomization, blinding, and placebo administration. The percentage of total publications (n=228) for each category receiving a Jadad score is presented in Table 3 below.

**Table 3:**
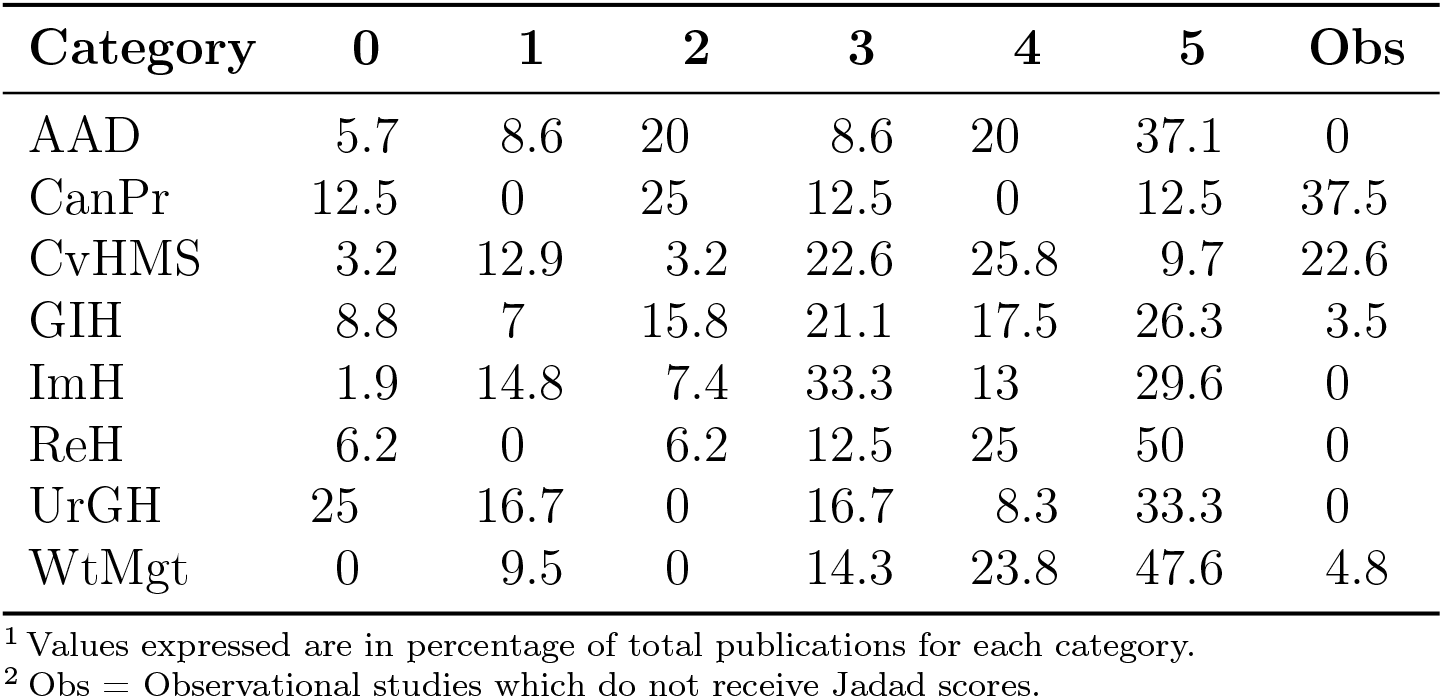
Percentage counts of studies in each health category, grouped by a Jadad scores from 0 (very poor) to 5 (rigorous).

From Table 3, it can be observed that most studies in AAD, ReH, and WtMgt were of high quality with regards to adherence to employment of controls and randomisation. The corresponding quality of evidence for claims in these categories is expected to be high. In CanPr, the majority of the studies were either Observational, or had low Jadad scores. Studies relating to CvHMS, ImH and UrGH included many publications in mid-Jadad scores (Scores 2 and 3). This suggests that future studies investigating health claims in these categories may require the greater use of placebo controls, blinding steps and better randomisation protocols. The distribution of Jadad scores for the selected publications each year is presented in **Supplementary Figure 10**.

#### Randomisation, comparators, and bias controls

Placebo controlled studies are an effective way of providing a comparator for observed changes. Among the 282 cases, only 6 did not use placebo or comparator (two cases from [34], two cases from [35], one case from [36] and one case from [37]). There were 17 observational cases using low consumption population groups as comparators since placebos as such are not relevant in those cases. Two studies ([30] and [38]) failed to specify if a placebo was used. In total, 239 cases reported the use of a placebo control of which 27 cases failed to provide details of the placebo composition. In all, 212 cases reported the use of placebo control and provided full details of the placebo-composition.

Blinding is an effective means of mitigating experimenter and participant bias. The 17 observational cases notwithstanding, 39 cases were unblinded. In all, 227 cases had some form of blinding, of which 16 cases were single blinded (blinded only to participant), 209 cases were double-blinded (blinded to the participant as well the experimenter), and 2 cases represented in the same publication [39] were triple-blinded (blinded to the participants, the experimenter and an independent statistician).

Lastly, the randomisation stage is a critical step for reducing bias, and allow independent statistical comparison. The 17 observational cases notwithstanding, only 29 cases failed to use any randomisation methods to group their participants. The remaining 237 cases were randomised. Compliance: Across the 228 publications, 114 failed to mention any compliance checks. Among the remaining 114 publications, 64 (28.1%) provided a numeric value for participant compliance. The median compliance noted was 97.0% (from 90.0 to 99.3% at probability-range of 0.25 to 0.75. Min-to-max is from 60 to 102%).

#### Funding sources and conflicts of interest (CoI)

The funding sources were grouped into charities/ boards with independent funding, universities, governments, companies/ corporations, and joint ventures across any of the parties.

Across the 228 publications, 31 make no mention of their funding sources, nor potential conflicts of interest by the authors. Based on **Supplementary Figure 8**, private companies/ corporations remain the dominant source of funding for research in the field of live dietary microbes. Nearly half (49.1%) of the publications reported some form of potential conflict of interest (CoI) by at least one author either via the receipt of consultation fees, grants to travel and speak in seminars on behalf of the company, being named in patents associated to their strain of interest, or being a sitting member of the funding body or board of research, share ownership or being the founder of the company. The only source of funding without any CoIs noted in the literature collection herein was in government research. The number of studies reporting positive outcomes in publications with declared CoIs is 63.6%, compared to those that have no CoIs to declare (36.4%).

#### Recruitment rates

Across the 228 publications, 101 do not provide any details on recruitment rates. In one study, the information was unclear [40], and 13 [23, 41, 42, 43, 44, 45, 46, 31, 47, 48, 49, 50, 51] were observational studies. Thus in all, information on recruitment rates were available for only 113 (49.6%) publications.

### 3.9 Research endpoints made in the publication

Across the literature, 64 unique research endpoints were recorded as primary outcomes. Their case counts of the endpoints coloured by the Jadad score of the corresponding publication are shown in Figure 6.

**Figure 6:**
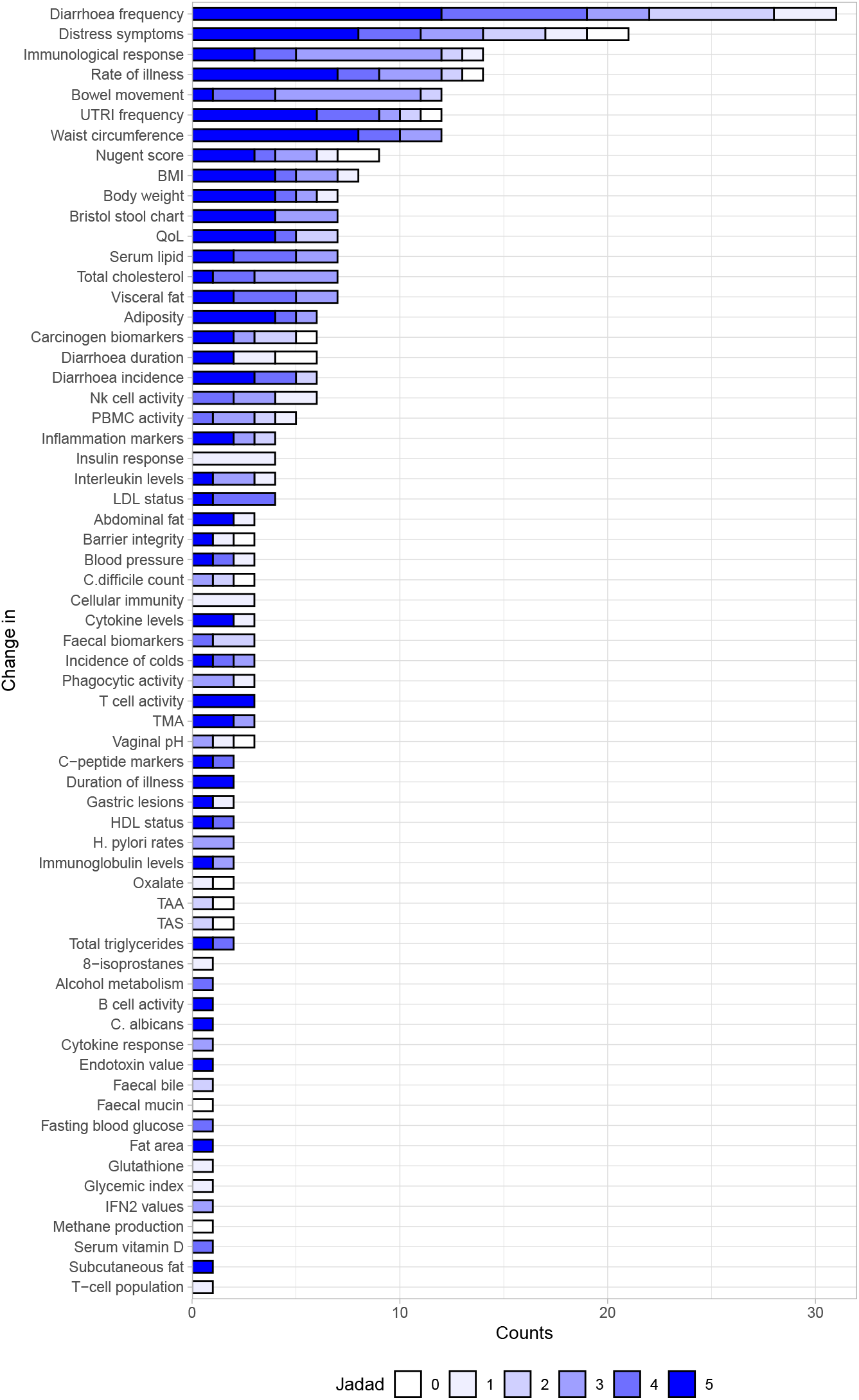
A stacked barplot of the counts of each endpoint made in the 228 publications recovered from the search protocol. The stacks are coloured according to the Jadad score.

For the research endpoints focusing on cellular immunity, total antioxidant status (TAS), total antioxidant activity (TAA), oxalate, colonic methane production, glycemic index, glutathione levels, interferon type II (IFN2) levels, faecal mucin, faecal bile, cytokine response, and 8-isoprostane levels only had evidence quality lower than Jadad 4.

## 4 Discussion

A wide variety of research findings, food products, methodologies and analyses have been presented across the collected literature that contribute to the evidence relating to the effectiveness of dietary microbes in maintaining general health and well-being in non-patient populations in various health categories.

### 4.1 Potential research gaps

Despite the impressive breadth of the collected evidence on the health benefits of dietary microbes, the following key research gaps have been identified:

#### Paucity of research

Paucity of research on traditional fermented foods, certain microbial genera, certain health categories, and research endpoints were noted.

Research was highly focused on the lactobacilli. Studies involving other microbes such as *Lactococcus*, *Pediococcus*, *Propionibacterium*, and *Weissella* were relatively rare.

Furthermore, despite the broad global variety of traditional fermented foods, only a relatively narrow range of products were recovered in our search. Many foods that are not produced on a global scale (such as kimchi [52], dêguê [53], filmjölk or långfil [14]) have been associated with health benefits, and a closer inspection of these foods may reveal novel and potentially beneficial microbes.

Lastly, the health categories chosen for study in this review were not equally represented, with UrGH and CanPr having fewer publications addressing the research question and satisfying the inclusion/exclusion criteria. Moreover, the quality of evidence for research endpoints such as vaginal pH, or faecal alfatoxin biomarkers was backed by low Jadad score publications.

#### Limited population representation

In almost all publications, the study population was recruited from the countries where the research institution was located. In **Supplementary Figure 9**, a connection-graph shows that most of the publications were from Europe while only two publications were from Africa.

The highest human genetic diversity is observed in the population of the African continent [54], and yet these populations are under-represented in research relating to the consumption of dietary microbes. Collaboration with countries from Africa and other under-represented regions such Oceania, particularly Pacific island communities, will provide a better representation of the health benefits associated with the consumption of dietary microbes across humanity, while also bringing more traditional fermented foods to global attention. Lastly, through collaborations, the scientific community would likely benefit from the knowledge of local researchers in these regions.

#### Problems with reporting data

There are some short-comings in the way results and outcomes are reported in publications. Significant inconsistency was noted across the publications with respect to the parameters measured at baseline. Base anthropometrics such as age, sex and BMI were often not reported. Furthermore, for studies involving dietary microbes, parameters such as faecal calprotectin, faecal mucin and faecal IgE were used as markers for intestinal distress. It may be worth including validating these endpoints and incorporating them as routine biomarkers required for dietary microbe based interventions.

The application of correct statistical methods and sample size calculations was also inconsistent. Most publications failed to mention base assumptions on the data distribution before selecting the statistical tools for hypothesis testing. In many instances, important outcomes were provided in the form of figures/graphs rather than a table with discrete values. This creates difficulties should there be a desire to include these outputs in meta-analyses. Moreover, the routine use of libre software to process the data and uploading relevant files on open-share repositories would significantly enhance research transparency and aid third-party meta-analyses and other study follow-ups.

Lastly, implementation of sample size calculations and the corresponding endpoints tested was inconsistent across the literature. In some publications many endpoints were tested on the basis of a single proxy-parameter used for sample size calculation. Moreover, the study designs were not robustly applied given that the median Jadad score across all publications was 3. More studies with high-quality designs are required to test the impact of live dietary microbes on health.

Other aspects such as the use of proprietary software, lack of compliance check reporting, and lack of adverse events reporting, significantly hamper further extrapolation.

### 4.2 Limitations of present study

#### Implied assumptions through aggregation

Many assumptions were made in this work by virtue of aggregation or collective summarisation. The outcomes were reported across the Jadad score range, experimental designs, sexes, age groups, and diversity of the microbes in the constituent food product. Although categories and groups were recognised during summarisation, the impact of food matrix effects on the final measured outcome could not be ascertained. Thus jointly reporting corresponding medians may fail to highlight the nuances in the data.

#### Adequate Intake (AI) value was observational

The adequate intake value noted here was an aggregated observation based on the reported frequency of a dose associated with each outcome. As mentioned in the preceding subsection, this makes assumptions relating to the comparability of each data point. Further work is required to define a statistical environment agnostic to experimental designs, sex / gender and age, and food types from which a limited and qualified statement of minimal effect doses can be elucidated using effect sizes.

#### Ambiguous use gender and sex

Inaccuracies in population characteristics were introduced as many publications used “sex” and “gender” interchangeably. A publication Kaufmann *et al*. [55] was included that conducted an intervention using a probiotic product on transwomen (male to female). We used sex and gender interchangeably owing to a lack of information in the selected publications. In the case of Kaufmann *et al*., participants were grouped under “female” since the intervention was performed to test the effect of probiotics on the pH and Nugent scores of neovaginal environments. Future research work will need to account for; and distinguish, gender identities from biological sex when reporting corresponding outcomes [56, 57].

## 5 Conclusion

Studies investigating the outcomes associated with dietary microbes consumed either as part of a fermented food or a probiotic in a non-patient population is diverse in terms of experimental designs, evidence quality, and constituent microbes. While some aspects have been well characterised with multiple studies of high quality backing the research claims, many others remain poorly studied, particularly with regards to cancer prevention and urogenital health.

Elucidating the beneficial effects of regular consumption of dietary microbes as part of a regular healthy lifestyle is challenging owing to the various unmeasured errors and confounders that are introduced anthropogenically or experimentally. Median intake value of 2 × 10^9^ CFU·day^−1^ of dietary microbes was associated with non-negative outcomes. Furthermore, older population groups with a median age of 39 years, were associated with positive outcomes.

The use of open data-sharing policies and libre software by primary researchers can significantly improve transparency, reproducibility and general quality of statistical analyses. Studied population requires greater representation in terms of ethnic, cultural and economic backgrounds, as they are expected to significantly influence health outcomes. Lastly, more high quality research is required to establish relevance of dietary microbes in maintaining general health. Validating and incorporating the measurement of faecal biomarkers (bile, mucin, calprotectin) at the baseline and endpoint may help integrate a more concrete measurement of potential adverse reaction to intervention.

## Funding source and conflicts of interest

This work was supported by the Institute for the Advancement of Food and Nutrition Sciences (IAFNS). IAFNS is a nonprofit science organization that pools funding from industry and advances science through the in-kind and financial contributions from private and public sector members.

Research in the Cotter laboratory is also funded by the European Union’s Horizon 2020 research and innovation programme, under the MASTER project [grant number 818368], by Science Foundation Ireland (SFI) under [grant number SFI/12/RC/2273_P2] (APC Microbiome Ireland), by SFI together with the Irish Department of Agriculture, Food and the Marine under [grant number SFI/16/RC/3835] (VistaMilk) and by Enterprise Ireland and industry in the Food for Health Ireland (FHI)-3 project, under [grant number TC/2018/0025]. Other authors have no other conflicts to declare.

## Data Availability

All data produced are available online at: https://osf.io/kvhe7/

## Acknowledgements

The authors would like to thank Mary Ellen Sanders for her valuable comments on the manuscript.

## Supporting data

**Supplementary Table 1:** File name is `Supplementary_Table1.csv`, which is the master-table of the data collected from the 282 publications selected here in.

**Supplementary Table 2:** File name is `Supplementary_Table2.csv`, which is the list of the reported microbial species in the literature and their corresponding updated name as on 8 February 2023.

**Supplementary Material:** File name is `Supplementary_Material.pdf`, which is the file containing all the supplementary supporting texts, codes, and figures.

## Notes

### Summary of Updates

1. Two of the author names were corrected by adding appropriate accents. 2. Since .csv files cannot be uploaded here, they have been converted to .tsv files. They have now been uploaded as well. 3. The manuscript noted the inclusion of a Supplementary_Materials.docx file. The file was rewritten in LaTeX and is now appended as a .pdf file. Also corresponding Supplementary Figures have been uploaded separately.

