## Supplementary figures and images for "The impact of live dietary microbes on health: a scoping review"

### Supplementary_Figure1

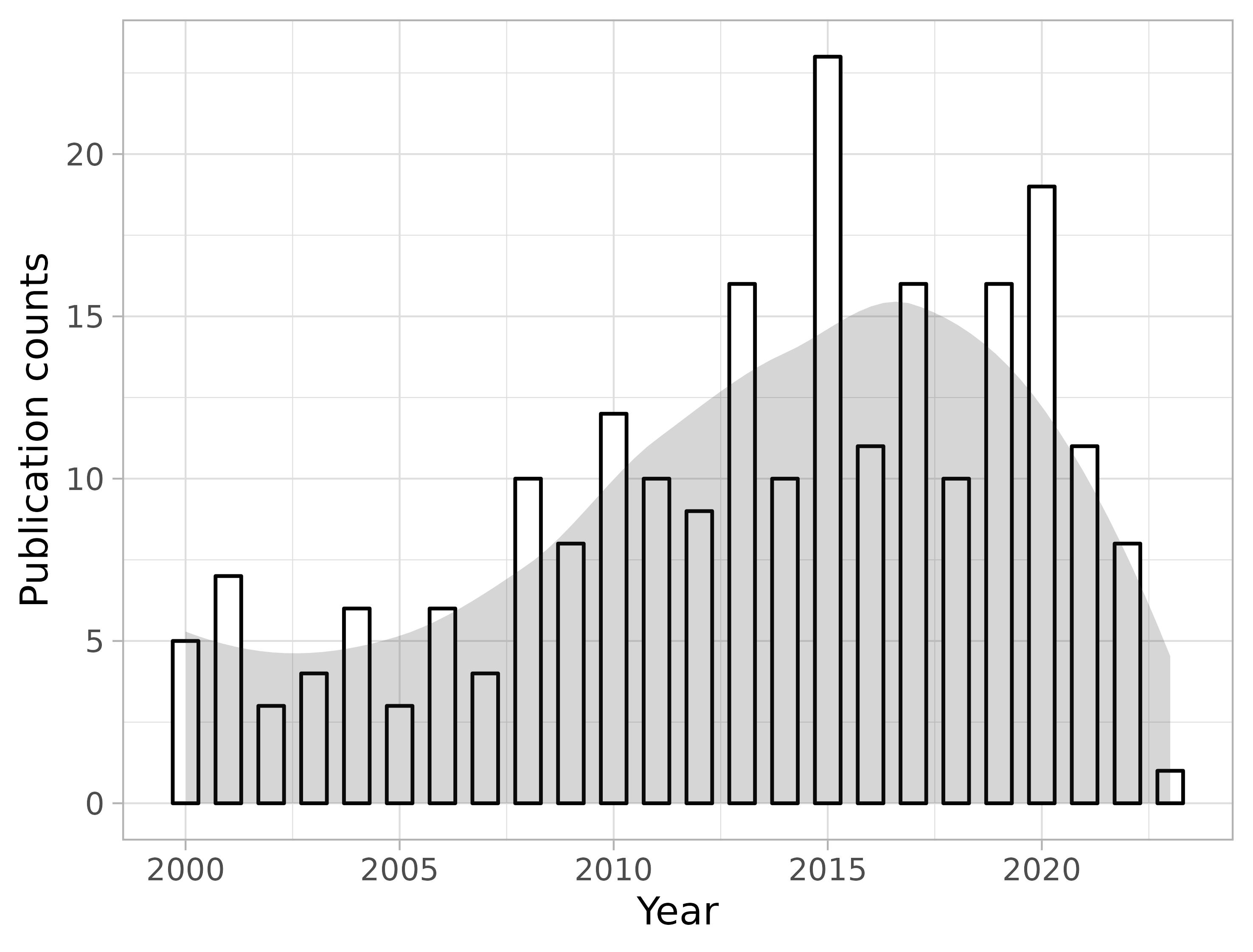

### Supplementary_Figure2

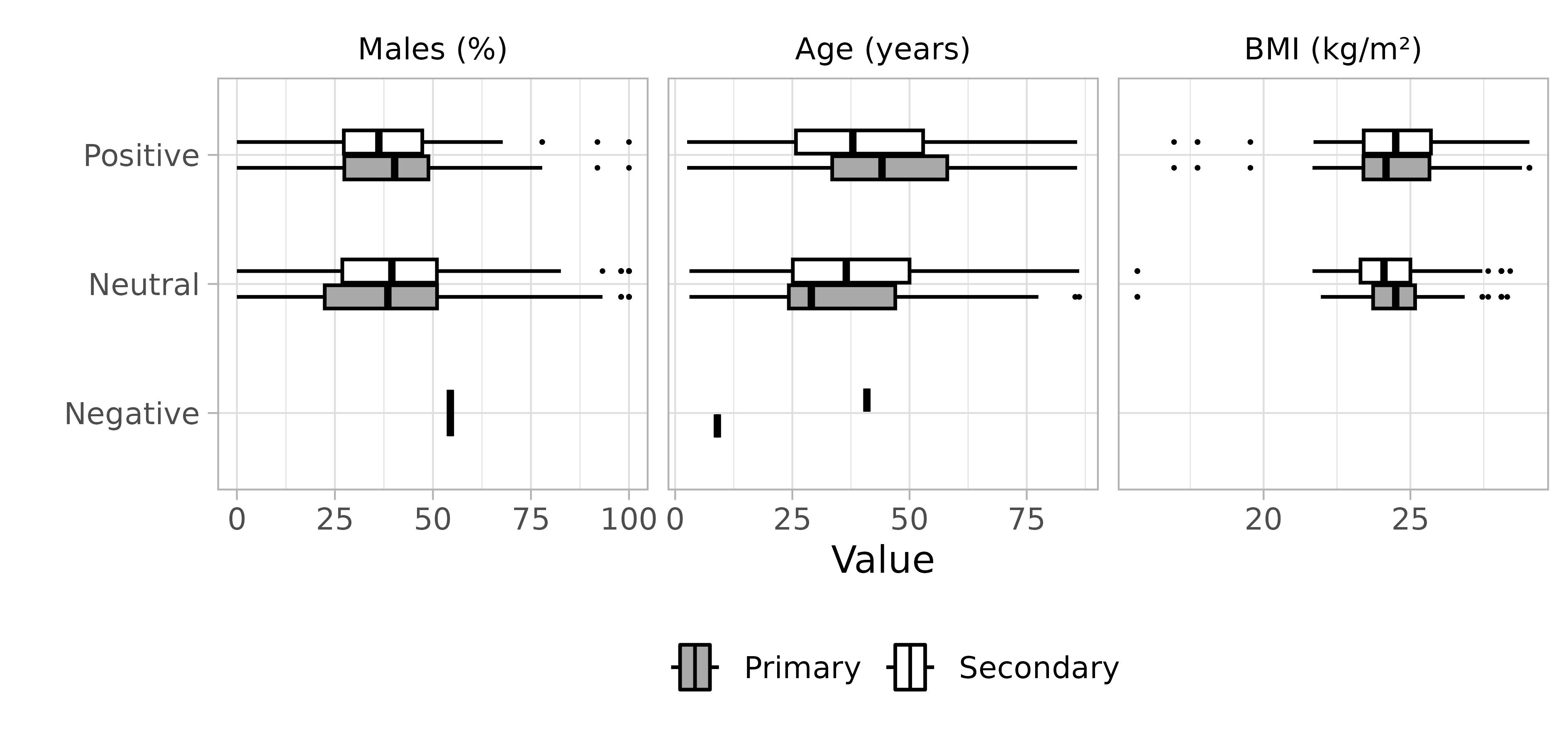

### Supplementary_Figure3

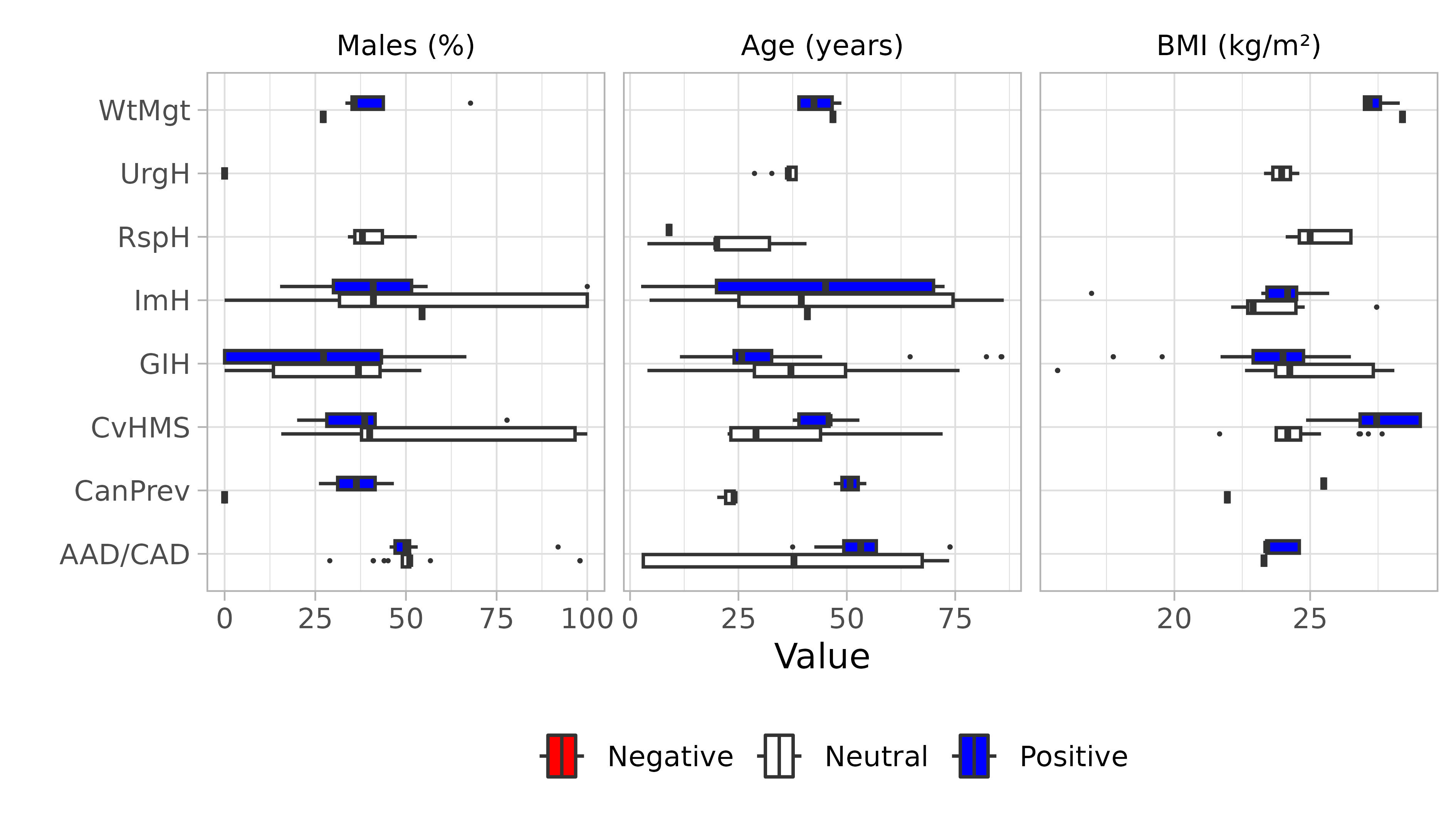

### Supplementary_Figure4

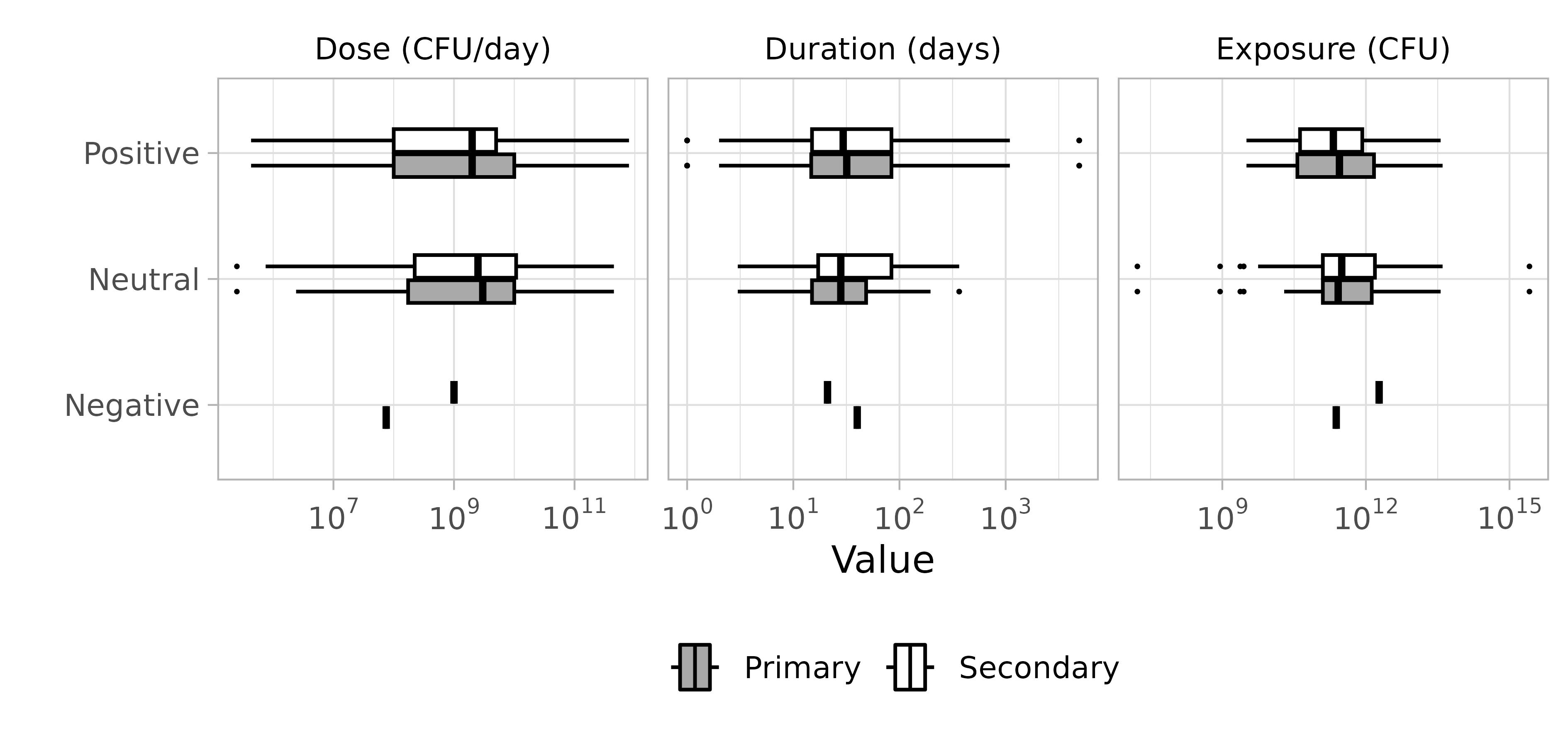

### Supplementary_Figure5

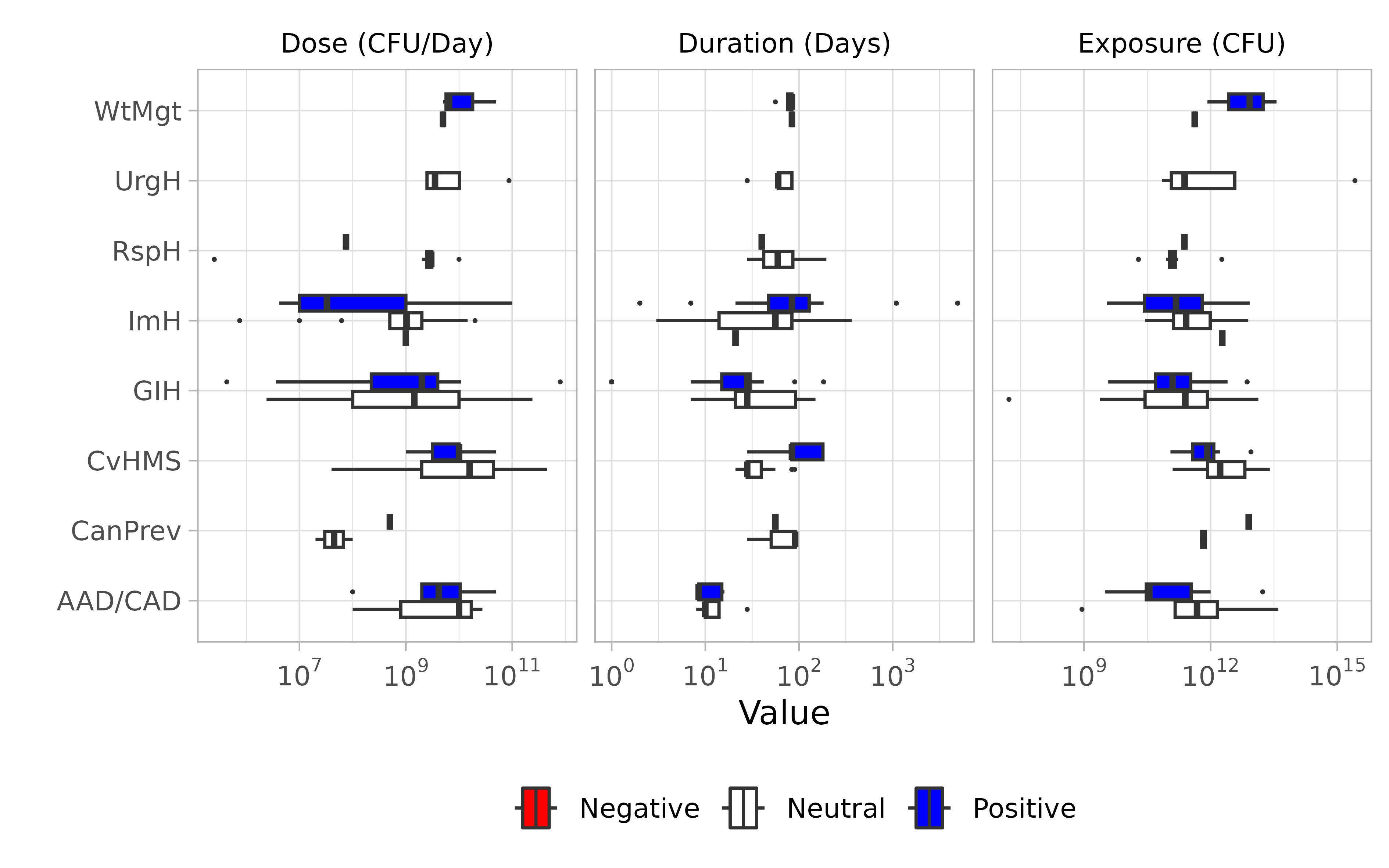

### Supplementary_Figure6

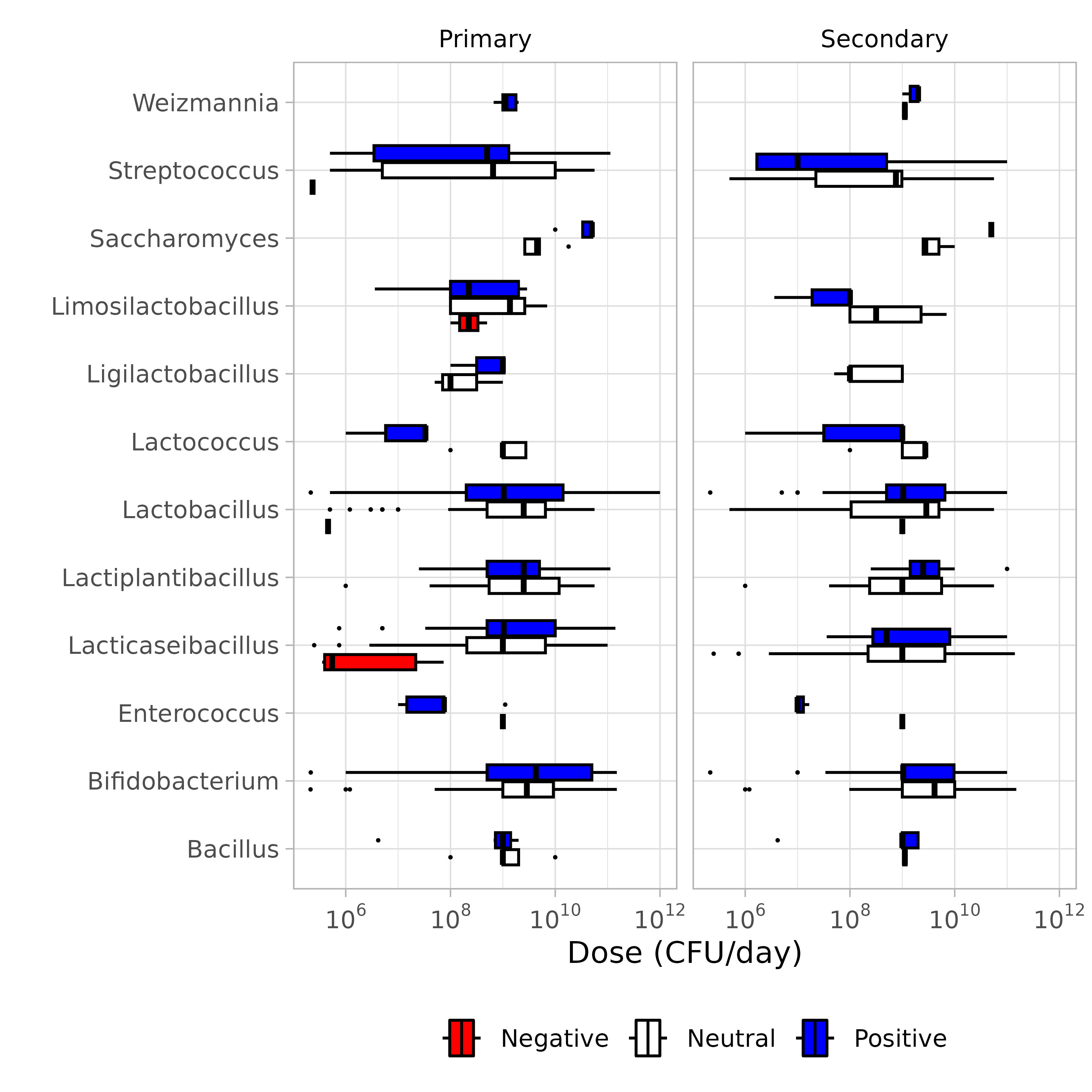

### Supplementary_Figure7

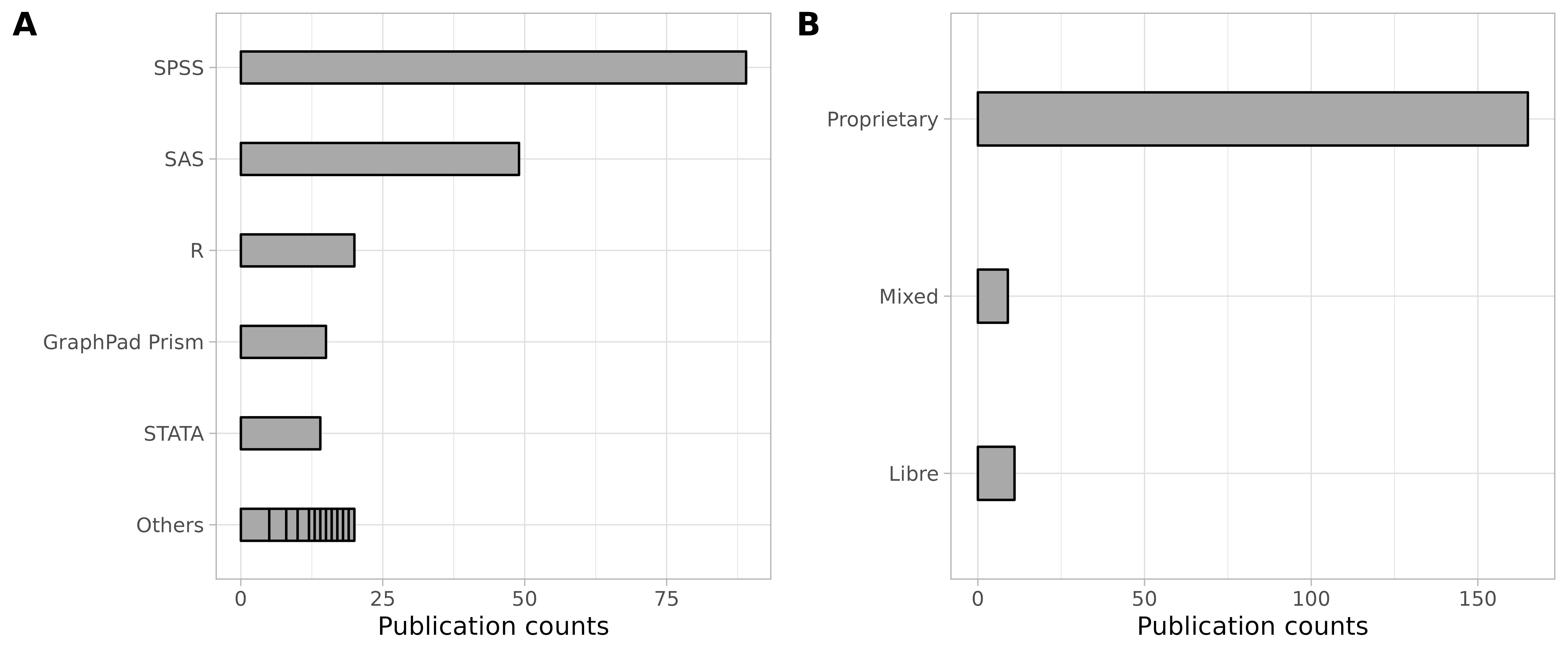

### Supplementary_Figure8

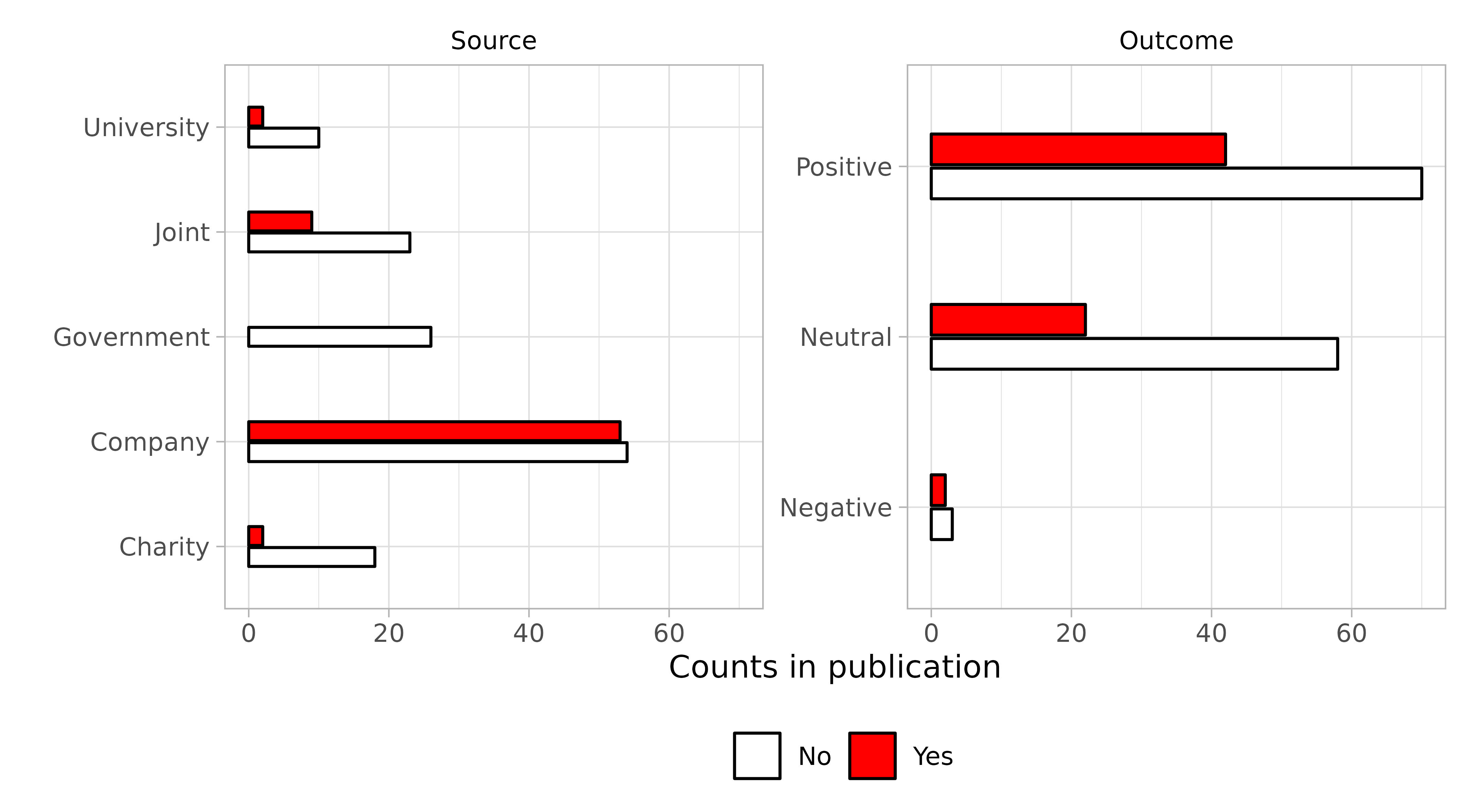

### Supplementary_Figure9

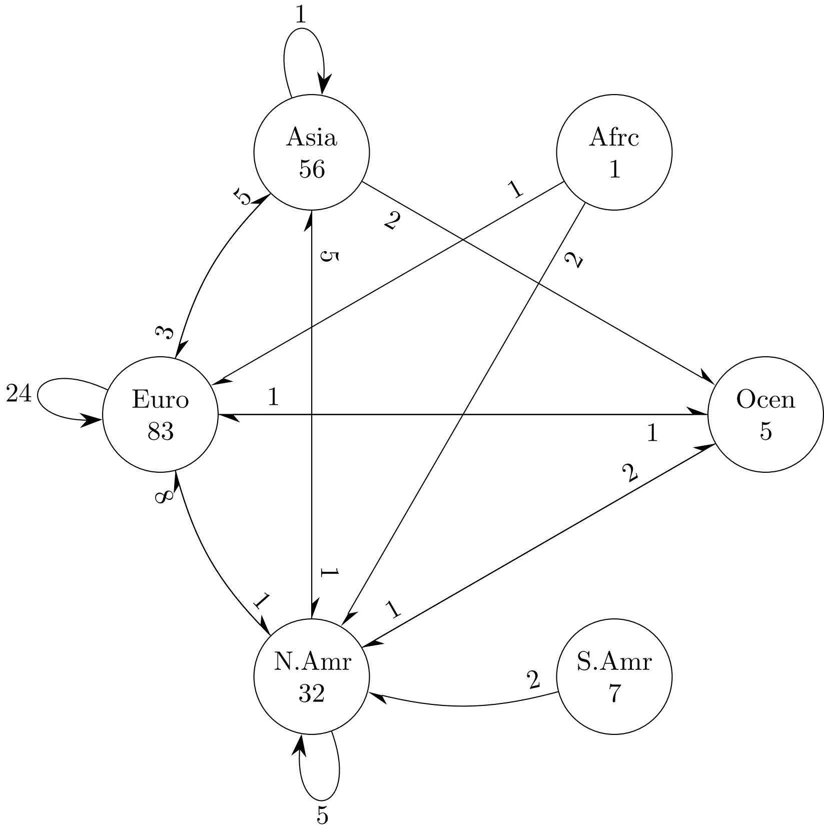

### Supplementary_Figure10

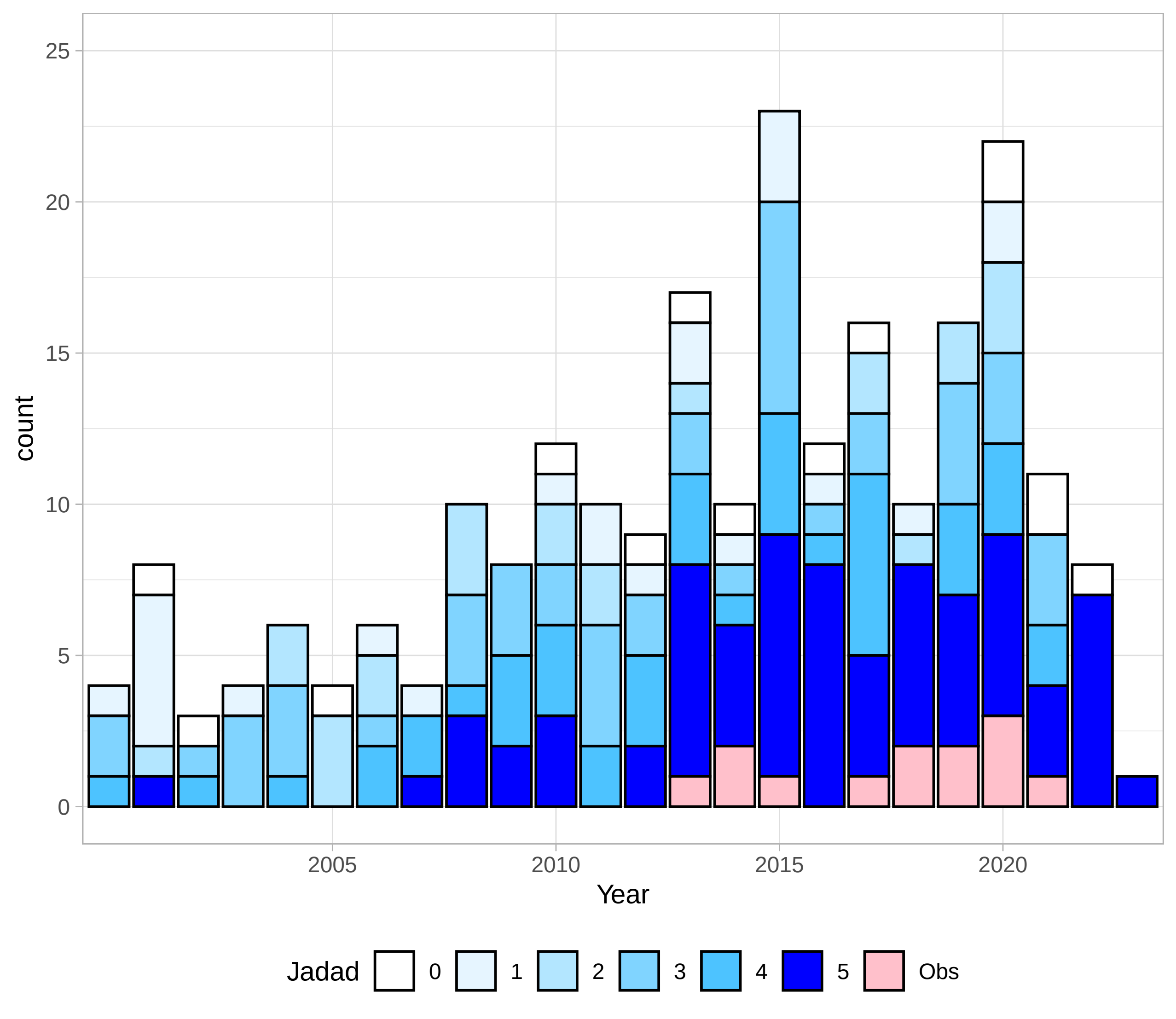
