## Supplementary Materials for "The impact of live dietary microbes on health: a scoping review"

[*]Shared first author
[1]Department of Food Biosciences, Teagasc Food Research Centre, Fermoy, Ireland.
[2]APC Microbiome Ireland, University College Cork, Cork, Ireland.
[3]Department of Biological Sciences, University of Limerick, Limerick, Ireland.
[4]Health Research Institute, University of Limerick, Limerick, Ireland.
[5]VistaMilk SFI Research Centre, Cork, Ireland.

Please find the pre-prints and supplementary files in our OSF repository: https://osf.io/kvhe7/.

### 1 Supplementary texts

#### 1.1 Search terms

For all searches, the first level for keywords describing the field was: `(probiot* OR nutr* OR ferm*)`. For the publication types, the terms used were: `(stud* OR trial*)`. Keywords within each level were combined with the `OR` operator, while the levels were combined with the `AND` operator.

Keywords for the specific category were:

**Antibiotic associated diarrhoea:** `(diarr* AND (antimic* OR antib*))`

**Gastrointestinal health:** `(gast* OR *intest*)`

**Immunological health:** `(immu* OR inflam*)`

**Cardiovascular health and metabolic syndrome:** `(cardio* OR diab* OR stroke* OR *glyce* OR *cholestero*)`

**Cancer:** `(cancer* OR carc* OR tumo* OR *neoplas* OR polyp* OR sarcom*)`

**Respiratory health:** `(resp* OR asth* OR pulmonary)`

**Weight management:** `(weigh* OR "BMI" OR obes* OR adipos*)`

**Urogenital health:** `(urogenital OR urin* OR vagi*)`

#### 1.2 Search structure

The following search term structure was adopted for each category:

```
(Keywords describing the field)
AND
(Keywords describing the publication types)
AND
(Keywords of the specific category)
```

The keyword structure was reformulated as an awk script. The results from the databases were saved as `.csv` files which were subsequently processed again using the awk filter on titles and abstracts. Next, the title and abstracts were screened to check for relevance based on predetermined exclusion criteria (Supplementary Code 1).

### 2 Awk code

The following awk code was used to quickly filter out irrelevant titles:

```
#!/bin/awk -f
BEGIN{ FS=","}
FNR==1 { print $0 > "terms.csv"  # write header row
         print $0 > "notterms.csv"    # to both files
         next
       }
       { name=$2
         sub(/^[ ]+|[ ]+$/,"",name)   # strip leading/trailing spaces from name
         if (name~/mice/ ||
          name~/wine/ ||
          name~/beer/ ||
          name~/chemo*/ ||
          name~/chicken/ ||
          name~/hen/ ||
          name~/spinal/||
          name~/depression/||
          name~/extact/ ||
          name~/stroke/ ||
          name~/chronic/ ||
          name~/drug/ ||
```

```
            name~/radiation/||
            name~/Parkinson/ ||
            name~/aller*/ ||
            name~/execise/ ||
            name~/preg*/ ||
            name~/infan*/ ||
            name~/feed/)
              print $0 > "terms.csv"
           else
              print $0 > "notterms.csv"
        }
END{}
```

Listing 1: awk code to filter results

The code generates a workfile `terms.csv` which contains titles with the irrelevant terms, and another workfile `notterms.csv` which contains the “cleaned” list of titles.

### 3 Supplementary figure

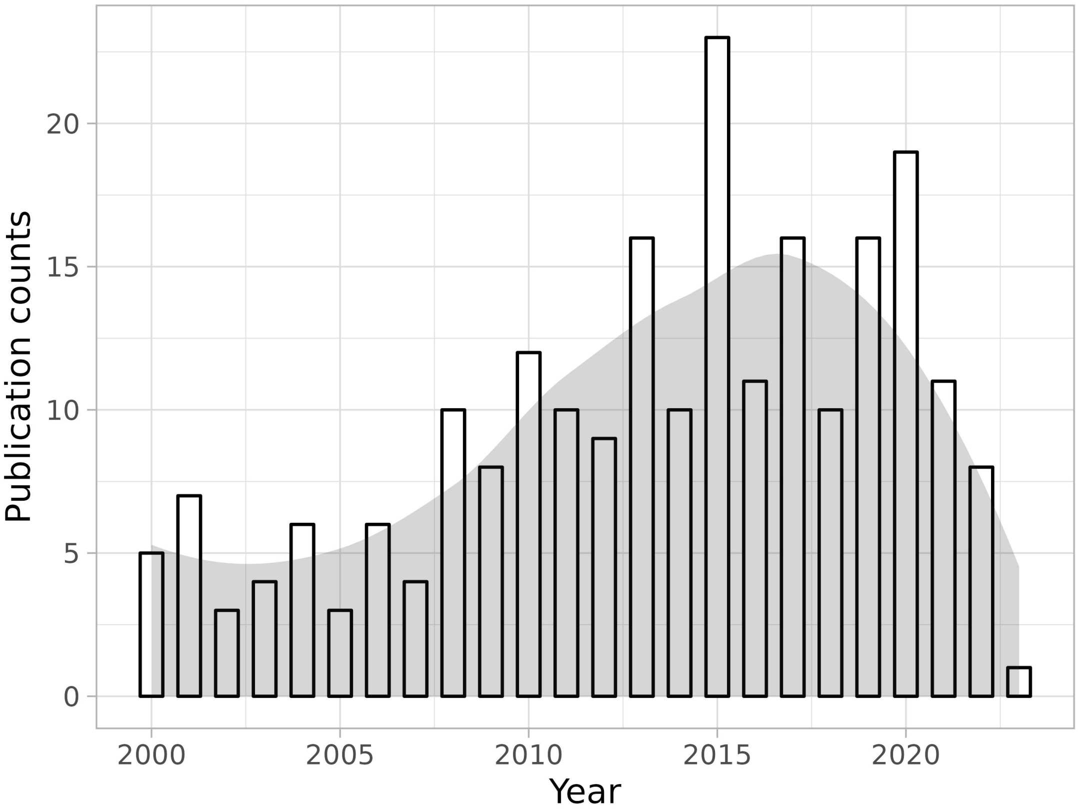


Supplementary Figure 1: Publication counts since the year 2000 to 2023 investigating consumption of dietary microbes via fermented food products. A loess-smoothened area curve was overlaid to understand the overall trend. Relevant publication peaked in 2016 to 2017.

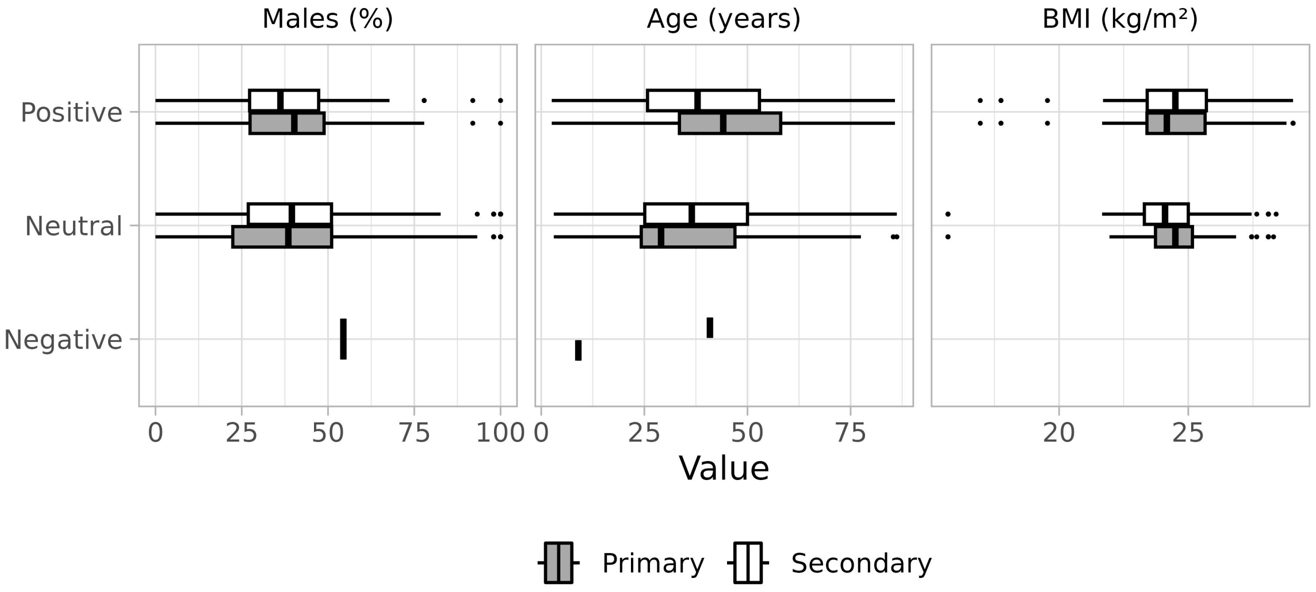


Supplementary Figure 2: The overall anthropometric measures of the studies administering dietary microbe rich foods grouped according study outcome (positive, neutral or negative). Males (%) represents the percentage of men in the study participation, Age (years) represents the mean age of the study participants, and BMI (kg·m$^{-2}$) represents the body-mass index of the participants.

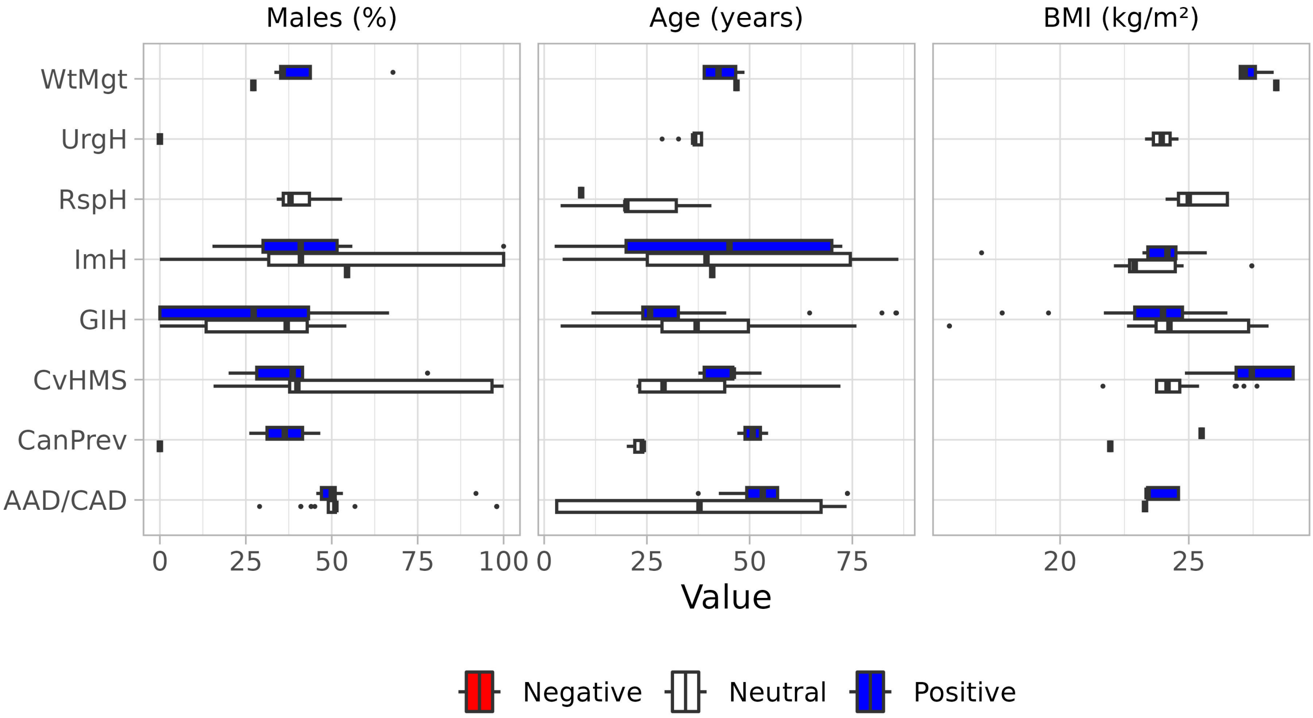


Supplementary Figure 3: The anthropometric measures of the experimental human studies administering dietary microbe rich foods grouped according to study secondary outcomes for each health category. Males (%) represents the percentage of men in the study participation, Age (years) represents the mean age of the study participants, and BMI (kg·m$^{-2}$) represents the body-mass index of the participants.

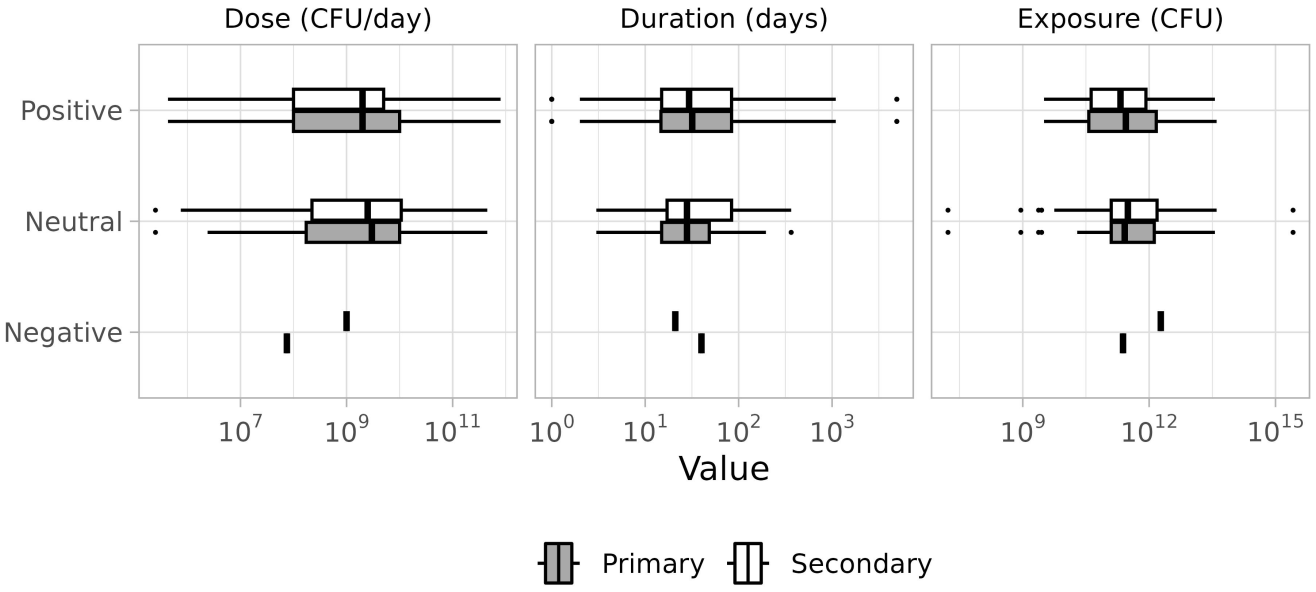


Supplementary Figure 4: Values of the study parameters grouped according to primary or secondary outcome. The first Panel "Dose" represents the microbial dose ingested by the volunteer each day. "Duration" represents the period of active consumption by the volunteer during the study. Exposure represents the total exposure to the dietary microbe by the study volunteer. It is calculated as the product of Dose and Duration.

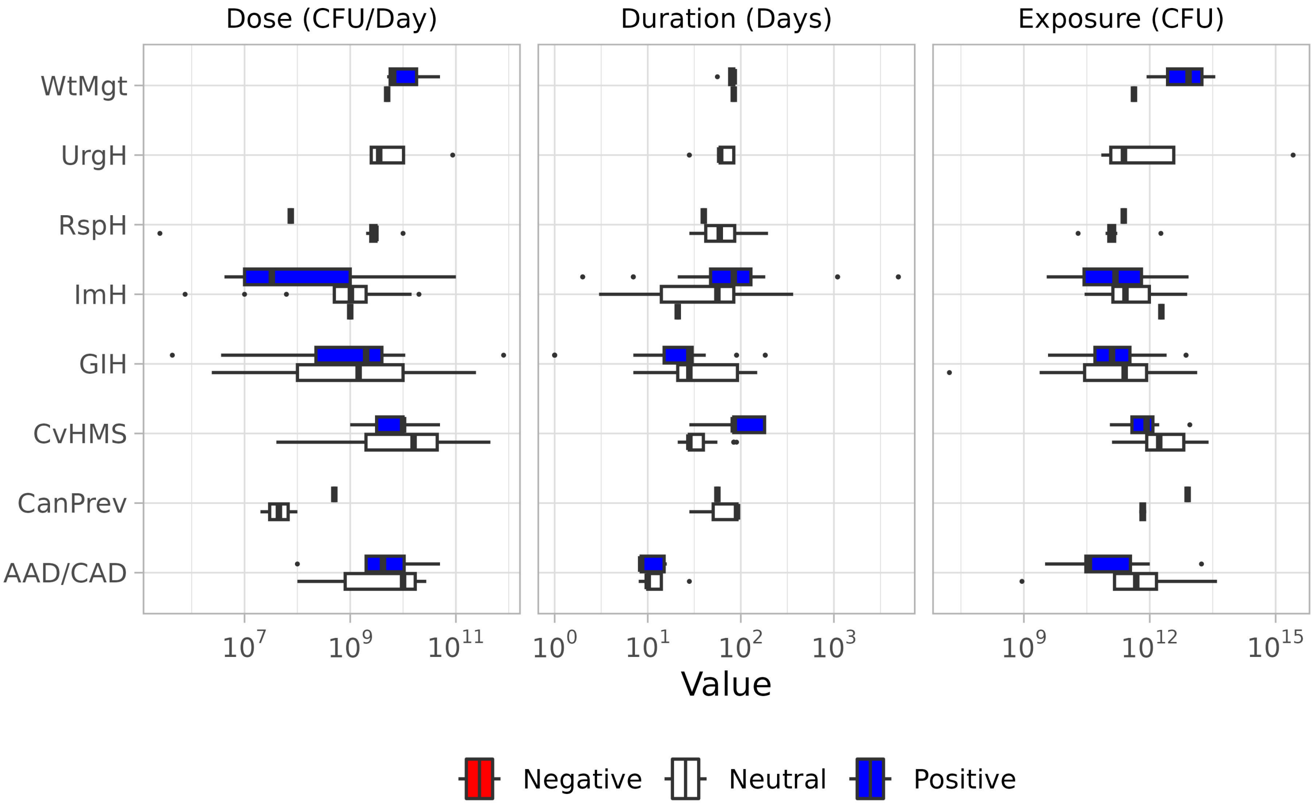


Supplementary Figure 5: Values of the study parameters grouped according to study outcome (negative, neutral and positive) for each health category. The first Panel "Dose" represents the microbial dose ingested by the volunteer each day. "Duration" represents the period of active consumption by the volunteer during the study. Exposure represents the total exposure to the dietary microbe by the study volunteer. It is calculated as the product of Dose and Duration.

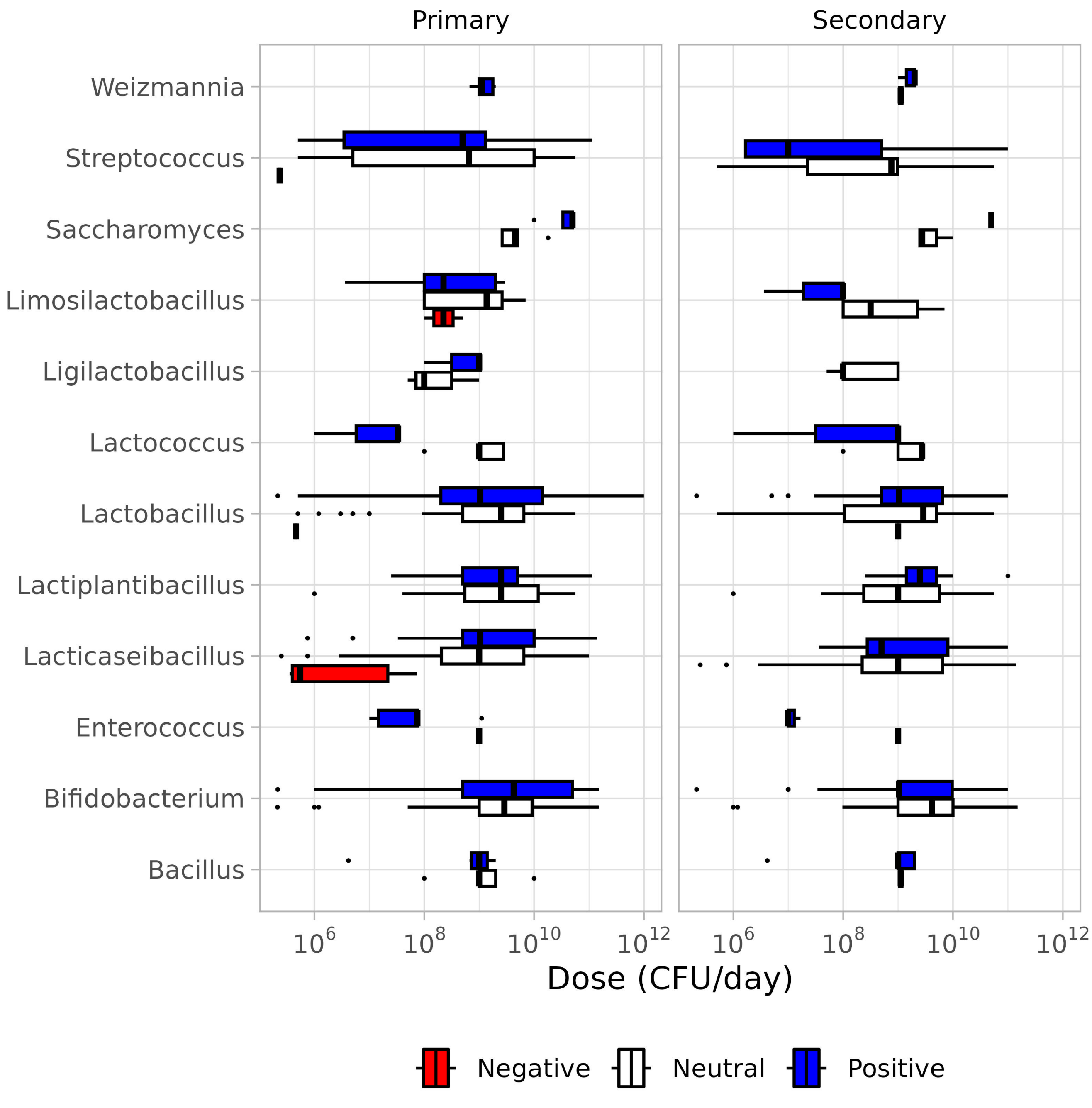


Supplementary Figure 6: Doses of each microbial genera reported in the fermented foods grouped according to study outcome (negative, neutral or positive) for Primary and Secondary claims.

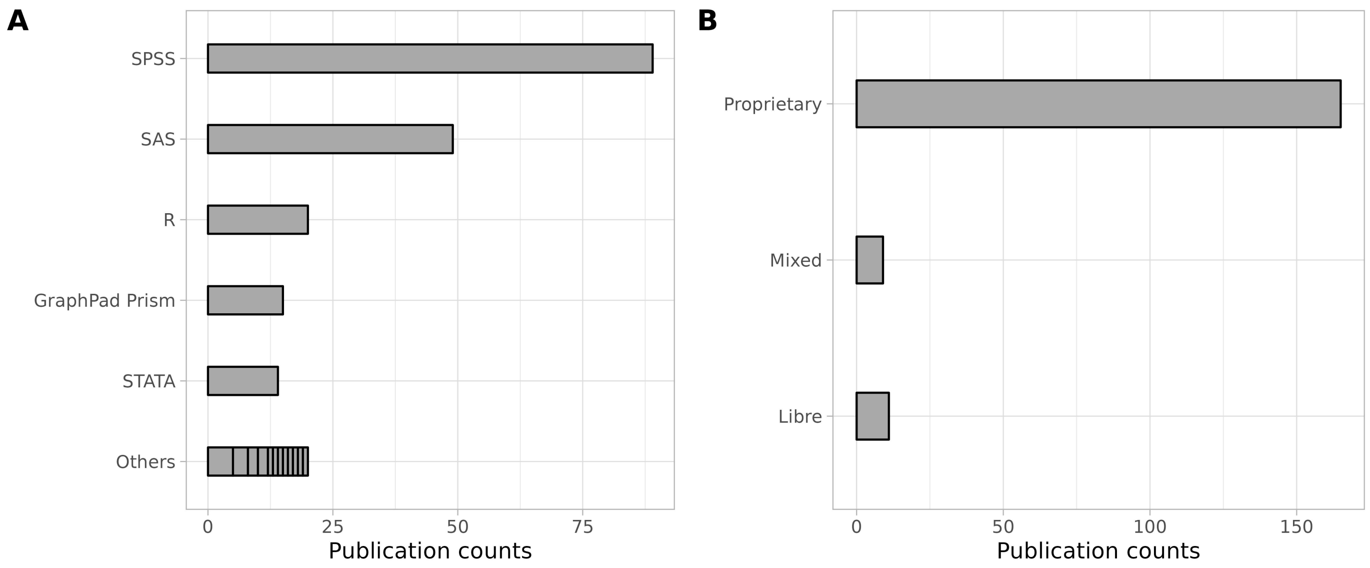


Supplementary Figure 7: Software and their code-sharing capability used in selected publications. Panel A represents the popularity (frequency of use) of the analytical software used. Panel B notes the number of times proprietary or Libre (and open source) software were used.

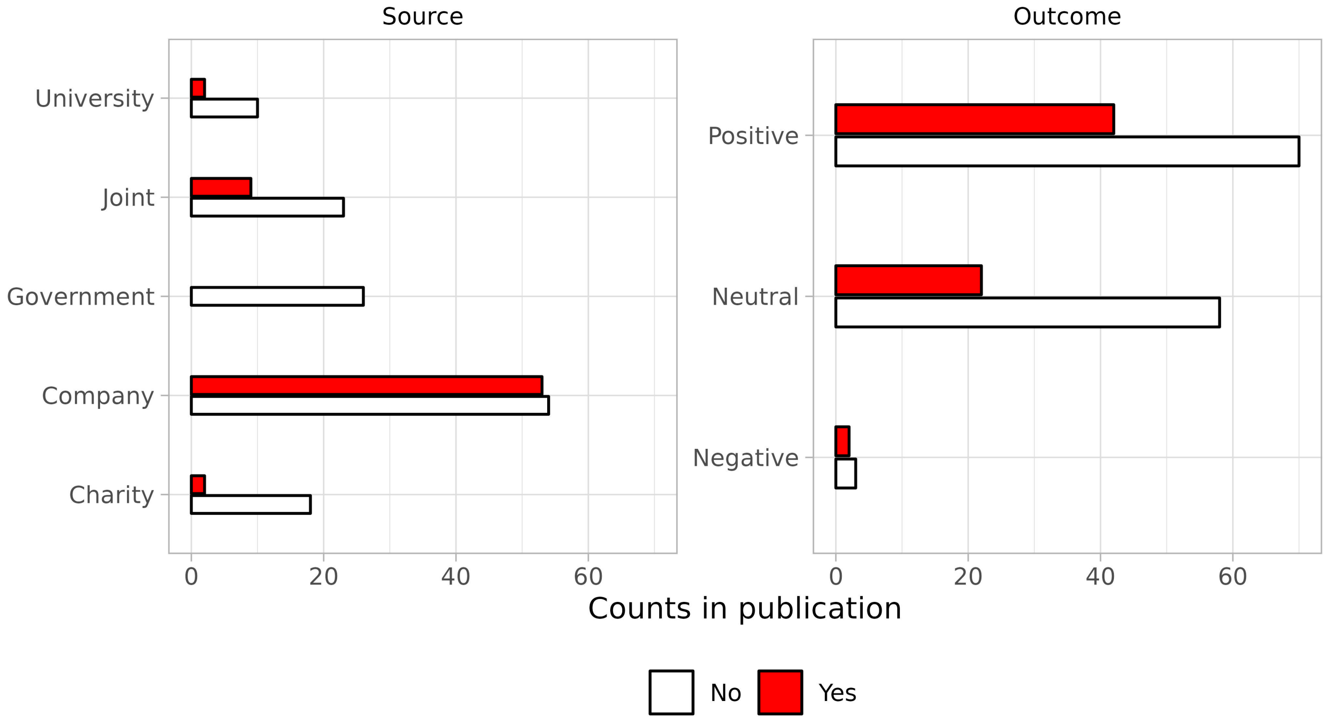


Supplementary Figure 8: Conflicts of interest noted in the selected publication grouped according to source of the funding (Panel A) or according to study outcome (Panel B). Filled colour represented either presence of a conflict of interest by at least one author (Yes, red) or no conflicts of interest by any author (No, white).

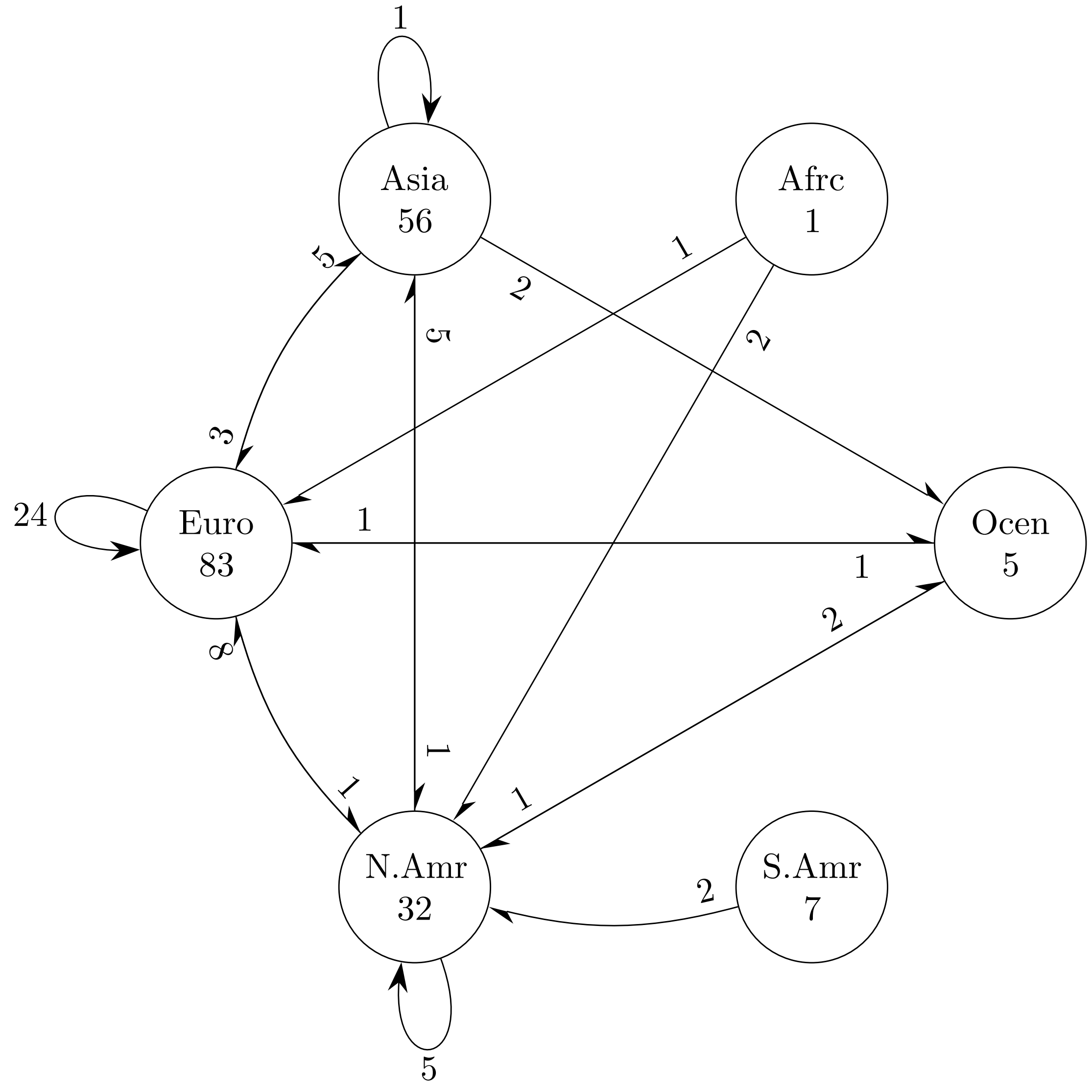


Supplementary Figure 9: Connections between continents where each research in each publication was performed. When a continent is at the root of an arrow, it was represented by the first author. A continent at the tip of the arrow is represented by co-authors. Looped arrows represent cases where the same continent was represented in the first as well as co-authors. Numbers along the path between two continents represent the number of connections between them. Values at the base of an arrow represents the number of connections the corresponding category has; as the primary outcome, with the category at the tip of the arrow as the secondary outcome.

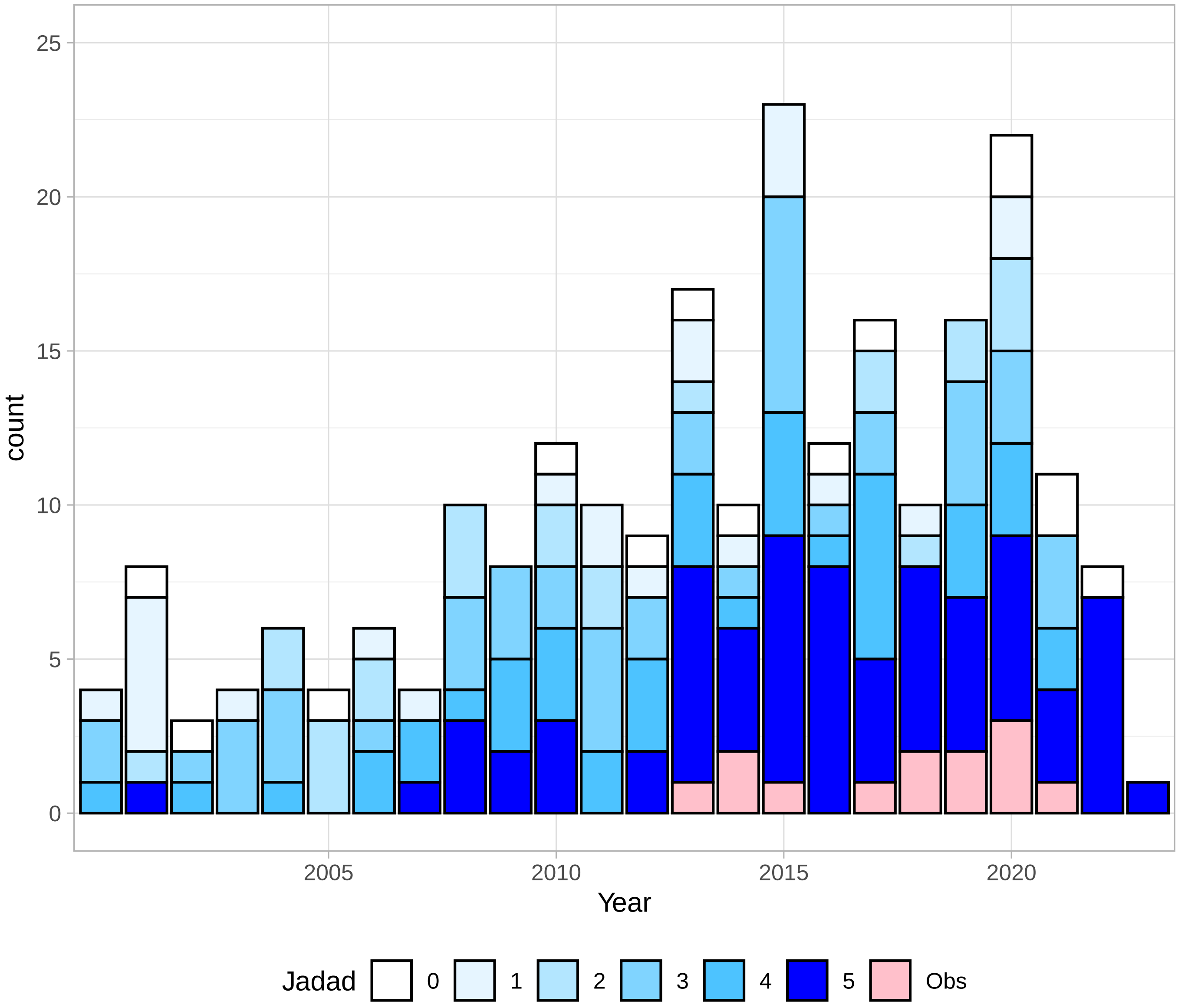


Supplementary Figure 10: Distribution of Publication according to Jadad scores.
